# Trends of Developmental Milestone Attainment in Israeli Children Between 2016-2020: A National Report

**DOI:** 10.1101/2023.02.05.23285482

**Authors:** Irena Girshovitz, Guy Amit, Inbal Goldshtein, Deena R. Zimmerman, Ravit Baruch, Pinchas Akiva, Meytal Avgil Tsadok, Yair Sadaka

**Affiliations:** KI Research Institute, Kfar Malal, Israel; Department of Maternal and Child Health, Public Health Division, Ministry of Health, Jerusalem, Israel; TIMNA Initiative, Big Data Platform, Israel Ministry of Health, Jerusalem, Israel; Neuro-Developmental Research Center, Mental Health Institute, Be’er-Sheva, Israel; Faculty of Health Sciences, Ben-Gurion University of the Negev, Be’er-Sheva, Israel

**Keywords:** Child development, Developmental surveillance, Trends, Israel, Milestone attainment

## Abstract

**Background:** The early years of children’s lives are critical for their healthy development. Although children’s growth and development rates may vary, a significant delay during early childhood could indicate a medical or a developmental disorder. Developmental surveillance is used worldwide by healthcare providers in routine encounters, as well as by educators and parents, to elicit concerns about child development. In this work, we used a national dataset of developmental assessments to describe temporal trends of milestone attainment rates and associations between milestone attainment and various sociodemographic factors.

**Methods:** The study included 1,002,700 children ages birth until 6 years with 4,441,689 developmental visits between the years 2016 and 2020. We used the Israeli developmental scale to assess the annual rates of failure to attain language, social and motoric milestones by the entire population, as well as by subgroups stratified by sociodemographic factors. We used multivariable logistic regression to analyze the impact of different sociodemographic factors on the odds of failure to attain milestones, while controlling for confounding.

**Results:** Milestone failure rates progressively increased over the examined years in all developmental domains, and most prominently in the language domain. Conversely, the rates of parental concern for developmental delay remained constant. In multivariable analysis, higher risk of milestone attainment failure was observed in children whose mothers were divorced, unemployed, immigrant, had lower education, of Bedouin origin or were over 40 years old when giving birth.

**Conclusions:** This report describes national trends of child development in the gross motor, fine motor, language, and social domains. An annual report of these trends may assist policy makers to objectively evaluate subgroups in need for intervention, and to assess the effectiveness of intervention programs in attempt to maximize the developmental potential of children in Israel.

## Background

Early childhood is considered an important period in the process of child development. According to recent studies, there has been a substantial increase in risk for growth and developmental delay over the last decade [1–3]. Although children’s growth and development rate may vary, a significant delay could indicate a medical or a developmental disorder. Early intervention has been proven to be critical in promoting language, cognitive, motor, socioemotional development, and educational success, alongside prevention of additional future developmental delays [4–11].

Developmental surveillance, based on milestone attainment, is used worldwide by pediatricians and healthcare providers during routine encounters, as well as by parents and educators to raise concern about child development.

The importance of developmental surveillance tools was recently stressed by the Centers for Disease Control and Prevention (CDC). In partnership with the American Academy of Pediatrics (AAP) they convened a panel of subject matter experts to identify a set of evidence-informed milestones for revision of the Learn the Signs and Act Early materials. These materials are commonly used for the practice of developmental surveillance [12].

Maternal child health clinics (MCHCs) are known in Israel as “Tipat Halav” (“drop of milk”)[13]. These free-of-charge clinics provide preventative care for children from aged from birth until 6 years. Parents are advised to visit the MCHC about a week after hospital discharge, and then at the age of 2, 4, 6, 9, 12, 18, 24, 36, 48 and 60 months [14]. In each MCHC visit, the child undergoes routine growth, physical and developmental evaluation by trained public health nurses, along with immunization according to the national immunization schedule.

Sociodemographic factors have been previously associated with developmental delay. Socioeconomic status [15–17], maternal age at time of child’s birth [15,16], sex of child [17], maternal marital status [16,18], cognitive stimulation of child [18,19] and number of child in the family [16] have been reported as being risk factors for developmental delay.

In this work, we use a national dataset to examine temporal trends and associations between different sociodemographic factors and milestone attainment rates.

Objective evaluation of the temporal trends in milestone attainment failure rates and its relations to sociodemographic factors may contribute to the understanding of social patterns in children’s healthcare. Furthermore, it may provide data-driven insights to stakeholders, assisting them to focus their efforts on specific subpopulations in need. If updated annually, this analysis may also be used to evaluate the effects of intervention programs.

## Methods

### Study population

The study population consists of all children visiting MCHCs between January 1^st^, 2016 and December 31^st^, 2020, including preterm children.

### Maternal child health clinics

All MCHCs are conducted according to Ministry of Health (MOH) national protocols [14]. The dataset used in this study contained an anonymized version of all data collected between 2016 and 2020 during visits to approximately 1000 MCHCs. These clinics are operated by the MOH, two municipalities (Jerusalem and Tel Aviv) and one health maintenance organization (Leumit), all of which use a designated electronic medical record (EMR) system named “Machshava Briah” (“Healthy Thought”) [14]. The database covers approximately 70% of the children from infancy to the age of 6 years, living in Israel [20]. It was previously utilized to construct population norms, referred as the Tipat Halav Israel Developmental Surveillance (THIS) Scale, based on over 3.7 million assessments of over 640 thousand children [20].

### Measured developmental outcomes

Threshold ages for each milestone were defined in accordance with THIS developmental scale [20]. For each developmental domain we calculated the percentage of children who failed to attain at least one milestone in a given year, where “Failure” was defined as inability to attain a milestone at an age equal to or greater than the THIS 95% threshold age.

In addition, we computed the proportion of children whose parents expressed concern regarding their development during the developmental evaluation in the MCHC, and the proportion of children whose developmental status was reported by the MCHC nurse as “inadequate for age”.

The following maternal factors were extracted: education, ethnic group, birth country, employment status, marital status and age at delivery (Table 1). In addition, we used the child’s age at milestone evaluation and the child’s sex. In some variables values were grouped, due to small numbers.

**Table 1.a.** Study population characteristics by year. In the religion variable, mothers whose origin was reported “Circassian”, “Other Muslim”, “Other Christian”, “Unknown” or “Other” were all sub-grouped as “Other”. In the maternal education level “Other” includes “Missing”, “No education” and “Studies in Yeshiva”. In the birth country variable, “Other” includes children whose mothers were reported as born in Europe, Other, Unknown, North America, Africa, Asia, South and Central America, Australia, and Oceania. In marital status, ‘Missing’, ‘widower’ and ‘other’ were reported as ‘Other’.

|  |  | Total number of Children |  |  |  |  |
| --- | --- | --- | --- | --- | --- | --- |
|  |  | 2016 | 2017 | 2018 | 2019 | 2020 |
| **n** |  | 422534 | 439369 | 456681 | 455942 | 394236 |
| **Maternal education, n (%)** | **Elementary** | 9530 | 9715 | 9784 | 9842 | 8107 |
|  | **High School** | 117661 | 121669 | 125123 | 122749 | 104013 |
|  | **Tertiary Education** | 35500 | 38634 | 42597 | 45359 | 42425 |
|  | **Academic** | 124969 | 133136 | 140590 | 142639 | 123922 |
|  | **Missing** | 134874 | 136215 | 138587 | 135353 | 115769 |
| **Ethnic group, n (%)** | **Christian Arab** | 6241 | 6391 | 6545 | 6375 | 5374 |
|  | **Druze** | 8621 | 9012 | 9180 | 9129 | 7547 |
|  | **Jewish** | 248900 | 257901 | 266650 | 269929 | 237924 |
|  | **Muslim Arab** | 94202 | 99691 | 102199 | 99663 | 84921 |
|  | **Muslim Bedouin** | 1177 | 1917 | 3735 | 6202 | 7996 |
|  | **Other** | 9437 | 9730 | 10170 | 10318 | 8912 |
|  | **Missing** | 53956 | 54727 | 58202 | 54326 | 41562 |
| **Sex of child, n (%)** | **Female** | 205548 | 213675 | 222102 | 221234 | 191276 |
|  | **Male** | 216986 | 225694 | 234579 | 234708 | 202960 |
| **Maternal birth country, n (%)** | **Ethiopia** | 6039 | 6323 | 6913 | 7069 | 6266 |
|  | **FSU** | 13867 | 15095 | 16488 | 16763 | 14079 |
|  | **Israel** | 344601 | 361705 | 373029 | 370504 | 319928 |
|  | **Other** | 25293 | 25239 | 25326 | 25883 | 23835 |
| **Maternal employment status, n (%)** | **Employed** | 165974 | 178179 | 195345 | 205890 | 189838 |
|  | **Unemployed** | 93411 | 95953 | 97224 | 94809 | 79416 |
|  | **Student** | 15608 | 17519 | 19318 | 20128 | 18225 |
|  | **Other** | 147541 | 147718 | 144794 | 135115 | 106757 |
| **Maternal marital status, n (%)** | **Married** | 370702 | 387562 | 400797 | 397968 | 342344 |
|  | **Unmarried** | 12132 | 13181 | 14371 | 15259 | 13847 |
|  | **Divorced** | 5286 | 5244 | 5278 | 4860 | 3773 |
|  | **Other** | 34414 | 33382 | 36235 | 37855 | 34272 |
| **Maternal age, n (%)** | **<=20** | 18631 | 18821 | 18464 | 17512 | 14206 |
|  | **20-40** | 391600 | 406961 | 421384 | 418611 | 360138 |
|  | **>40** | 11831 | 12865 | 13929 | 14506 | 12984 |
| **Child’s age at time of estimation, n (%)** | **0-1** | 187783 | 192493 | 197007 | 197809 | 185019 |
|  | **1-2** | 169254 | 173707 | 181267 | 181784 | 166682 |
|  | **2-3** | 106652 | 109820 | 114959 | 115508 | 104789 |
|  | **3-4** | 45226 | 50475 | 54075 | 52957 | 23917 |
|  | **4-5** | 9602 | 10850 | 12000 | 11760 | 9968 |
|  | **5+** | 15888 | 18666 | 22194 | 21439 | 9337 |

**Table 1.b.** Study population by reports of parental and nurse’s concern regarding child development

|  |  | parental report of developmental concern |  |  |  |  | Nurse's report of developmental concern |  |  |  |  |
| --- | --- | --- | --- | --- | --- | --- | --- | --- | --- | --- | --- |
|  |  | 2016 | 2017 | 2018 | 2019 | 2020 | 2016 | 2017 | 2018 | 2019 | 2020 |
| **n** |  | 16683 | 16062 | 16236 | 16304 | 12940 | 16120 | 17619 | 19437 | 21085 | 16324 |
| **Maternal education, n (%)** | **Elementary** | 399 (4.2)* | 389 (4.0)* | 367 (3.8)* | 359 (3.6)* | 287 (3.5)* | 513 (5.4)* | 530 (5.5)* | 587 (6.0)* | 613 (6.2)* | 488 (6.0)* |
|  | **High School** | 5315 (4.5)* | 5229 (4.3)* | 5250 (4.2)* | 5210 (4.2)* | 4203 (4.0)* | 5568 (4.7)* | 6189 (5.1)* | 6763 (5.4)* | 7391 (6.0)* | 5690 (5.5)* |
|  | **Tertiary Education** | 1391 (3.9)* | 1312 (3.4)* | 1433 (3.4)* | 1507 (3.3)* | 1339 (3.2)* | 1299 (3.7)* | 1398 (3.6)* | 1700 (4.0)* | 1892 (4.2)* | 1649 (3.9)* |
|  | **Academic** | 4436 (3.5)* | 4236 (3.2)* | 4318 (3.1)* | 4280 (3.0)* | 3428 (2.8)* | 4113 (3.3)* | 4564 (3.4)* | 5026 (3.6)* | 5450 (3.8)* | 4177 (3.4)* |
|  | **Missing** | 5142 (3.8)* | 4896 (3.6)* | 4868 (3.5)* | 4948 (3.7)* | 3683 (3.2)* | 4627 (3.4)* | 4938 (3.6)* | 5361 (3.9)* | 5739 (4.2)* | 4320 (3.7)* |
| **Ethnic group, n (%)** | **Christian Arab** | 160 (2.6)* | 150 (2.3)* | 141 (2.2)* | 174 (2.7)* | 130 (2.4)* | 240 (3.8)* | 239 (3.7)* | 281 (4.3)* | 335 (5.3)* | 202 (3.8)* |
|  | **Druze** | 401 (4.7)* | 348 (3.9)* | 312 (3.4)* | 273 (3.0)* | 195 (2.6)* | 417 (4.8)* | 514 (5.7)* | 603 (6.6)* | 594 (6.5)* | 389 (5.2)* |
|  | **Jewish** | 9957 (4.0)* | 9403 (3.6)* | 9412 (3.5)* | 9477 (3.5)* | 7549 (3.2)* | 8691 (3.5)* | 9462 (3.7)* | 10164 (3.8)* | 11228 (4.2)* | 8938 (3.8)* |
|  | **Muslim Arab** | 3662 (3.9)* | 3851 (3.9)* | 3780 (3.7)* | 3869 (3.9)* | 3112 (3.7)* | 4330 (4.6)* | 4804 (4.8)* | 5323 (5.2)* | 5663 (5.7)* | 4334 (5.1)* |
|  | **Muslim Bedouin** | 14 (1.2)* | 19 (1.0)* | 40 (1.1)* | 83 (1.3)* | 142 (1.8)* | 25 (2.1)* | 50 (2.6)* | 114 (3.1)* | 217 (3.5)* | 278 (3.5)* |
|  | **Other** | 417 (4.4)* | 425 (4.4)* | 559 (5.5)* | 529 (5.1)* | 468 (5.3)* | 489 (5.2)* | 550 (5.7)* | 723 (7.1)* | 810 (7.9)* | 660 (7.4)* |
|  | **Missing** | 2072 (3.8)* | 1866 (3.4)* | 1992 (3.4)* | 1899 (3.5)* | 1344 (3.2)* | 1928 (3.6)* | 2000 (3.7)* | 2229 (3.8)* | 2238 (4.1)* | 1523 (3.7)* |
| **Sex of child, n (%)** | **Female** | 6286 (3.1)* | 6062 (2.8)* | 6010 (2.7)* | 5963 (2.7)* | 4733 (2.5)* | 6350 (3.1)* | 6985 (3.3)* | 7528 (3.4)* | 8178 (3.7)* | 6236 (3.3)* |
|  | **Male** | 10397 (4.8)* | 10000 (4.4)* | 10226 (4.4)* | 10341 (4.4)* | 8207 (4.0)* | 9770 (4.5)* | 10634 (4.7)* | 11909 (5.1)* | 12907 (5.5)* | 10088 (5.0)* |
| **Maternal birth country, n (%)** | **Ethiopia** | 213 (3.5)* | 223 (3.5)* | 246 (3.6)* | 275 (3.9)* | 228 (3.6)* | 315 (5.2)* | 378 (6.0)* | 409 (5.9)* | 549 (7.8)* | 453 (7.2)* |
|  | **FSU** | 641 (4.6)* | 648 (4.3)* | 780 (4.7)* | 825 (4.9)* | 710 (5.0)* | 677 (4.9)* | 821 (5.4)* | 1037 (6.3)* | 1154 (6.9)* | 985 (7.0)* |
|  | **Israel** | 13244 (3.8)* | 13093 (3.6)* | 12963 (3.5)* | 12967 (3.5)* | 10193 (3.2)* | 12876 (3.7)* | 14172 (3.9)* | 15383 (4.1)* | 16558 (4.5)* | 12649 (4.0)* |
|  | **Other** | 1161 (4.6)* | 993 (3.9)* | 1040 (4.1)* | 913 (3.5)* | 797 (3.3)* | 1020 (4.0)* | 1064 (4.2)* | 1184 (4.7)* | 1258 (4.9)* | 1063 (4.5)* |
| **Maternal employment status, n (%)** | **Employed** | 6353 (3.8)* | 6186 (3.5)* | 6549 (3.4)* | 6789 (3.3)* | 5778 (3.0)* | 6018 (3.6)* | 6611 (3.7)* | 7619 (3.9)* | 8745 (4.2)* | 7311 (3.9)* |
|  | **Unemployed** | 4126 (4.4)* | 3992 (4.2)* | 3972 (4.1)* | 4036 (4.3)* | 3209 (4.0)* | 4517 (4.8)* | 4996 (5.2)* | 5509 (5.7)* | 5847 (6.2)* | 4511 (5.7)* |
|  | **Student** | 530 (3.4)* | 574 (3.3)* | 591 (3.1)* | 611 (3.0)* | 547 (3.0)* | 490 (3.1)* | 521 (3.0)* | 710 (3.7)* | 799 (4.0)* | 635 (3.5)* |
|  | **Other** | 5674 (3.8)* | 5310 (3.6)* | 5124 (3.5)* | 4868 (3.6)* | 3406 (3.2)* | 5095 (3.5)* | 5491 (3.7)* | 5599 (3.9)* | 5694 (4.2)* | 3867 (3.6)* |
| **Maternal marital status, n (%)** | **Married** | 14432 (3.9)* | 14077 (3.6)* | 14057 (3.5)* | 14030 (3.5)* | 10979 (3.2)* | 13945 (3.8)* | 15347 (4.0)* | 16768 (4.2)* | 18106 (4.5)* | 13984 (4.1)* |
|  | **Unmarried** | 560 (4.6)* | 542 (4.1)* | 660 (4.6)* | 687 (4.5)* | 629 (4.5)* | 625 (5.2)* | 703 (5.3)* | 834 (5.8)* | 1014 (6.6)* | 818 (5.9)* |
|  | **Divorced** | 283 (5.4)* | 277 (5.3)* | 297 (5.6)* | 298 (6.1)* | 213 (5.6)* | 264 (5.0)* | 315 (6.0)* | 399 (7.6)* | 359 (7.4)* | 272 (7.2)* |
|  | **Other** | 1408 (4.1)* | 1166 (3.5)* | 1222 (3.4)* | 1289 (3.4)* | 1119 (3.3)* | 1286 (3.7)* | 1254 (3.8)* | 1436 (4.0)* | 1606 (4.2)* | 1250 (3.6)* |
| **Maternal age, n (%)** | **<=20** | 608 (3.3)* | 613 (3.3) | 574 (3.1)<sup>‡</sup> | 579 (3.3) | 445 (3.1)* | 669 (3.6)* | 770 (4.1)* | 762 (4.1)* | 891 (5.1)* | 603 (4.2)* |
|  | **20-40** | 15518 (4.0)* | 14878 (3.7) | 15041 (3.6)<sup>‡</sup> | 14985 (3.6) | 11763 (3.3)* | 14895 (3.8)* | 16181 (4.0)* | 17921 (4.3)* | 19222 (4.6)* | 14894 (4.1)* |
|  | **>40** | 540 (4.6)* | 553 (4.3) | 556 (4.0)<sup>‡</sup> | 607 (4.2) | 572 (4.4)* | 543 (4.6)* | 649 (5.0)* | 684 (4.9)* | 844 (5.8)* | 668 (5.1)* |
| **Child's age at time of estimation, n (%)** | **0-1** | 4536 (2.4)* | 4226 (2.2)* | 3976 (2.0)* | 3885 (2.0)* | 2730 (1.5)* | 5234 (2.8)* | 5682 (3.0)* | 5871 (3.0)* | 6239 (3.2)* | 4416 (2.4)* |
|  | **1-2** | 5031 (3.0)* | 4664 (2.7)* | 4592 (2.5)* | 4747 (2.6)* | 4132 (2.5)* | 4909 (2.9)* | 5233 (3.0)* | 5790 (3.2)* | 6483 (3.6)* | 5352 (3.2)* |
|  | **2-3** | 5315 (5.0)* | 4996 (4.5)* | 5197 (4.5)* | 5179 (4.5)* | 4842 (4.6)* | 4601 (4.3)* | 4828 (4.4)* | 5595 (4.9)* | 6149 (5.3)* | 5445 (5.2)* |
|  | **3-4** | 1950 (4.3)* | 2205 (4.4)* | 2315 (4.3)* | 2350 (4.4)* | 1216 (5.1)* | 1508 (3.3)* | 1941 (3.8)* | 2139 (4.0)* | 2357 (4.5)* | 1262 (5.3)* |
|  | **4-5** | 495 (5.2)* | 532 (4.9)* | 619 (5.2)* | 617 (5.2)* | 476 (4.8)* | 341 (3.6)* | 445 (4.1)* | 542 (4.5)* | 495 (4.2)* | 418 (4.2)* |
|  | **5+** | 445 (2.8)* | 512 (2.7)* | 640 (2.9)* | 561 (2.6)* | 269 (2.9)* | 373 (2.3)* | 530 (2.8)* | 687 (3.1)* | 698 (3.3)* | 323 (3.5)* |

\*P<0.001, ‡P<0.005 (Bonferroni adjusted p-value)

**Table 1.c.**
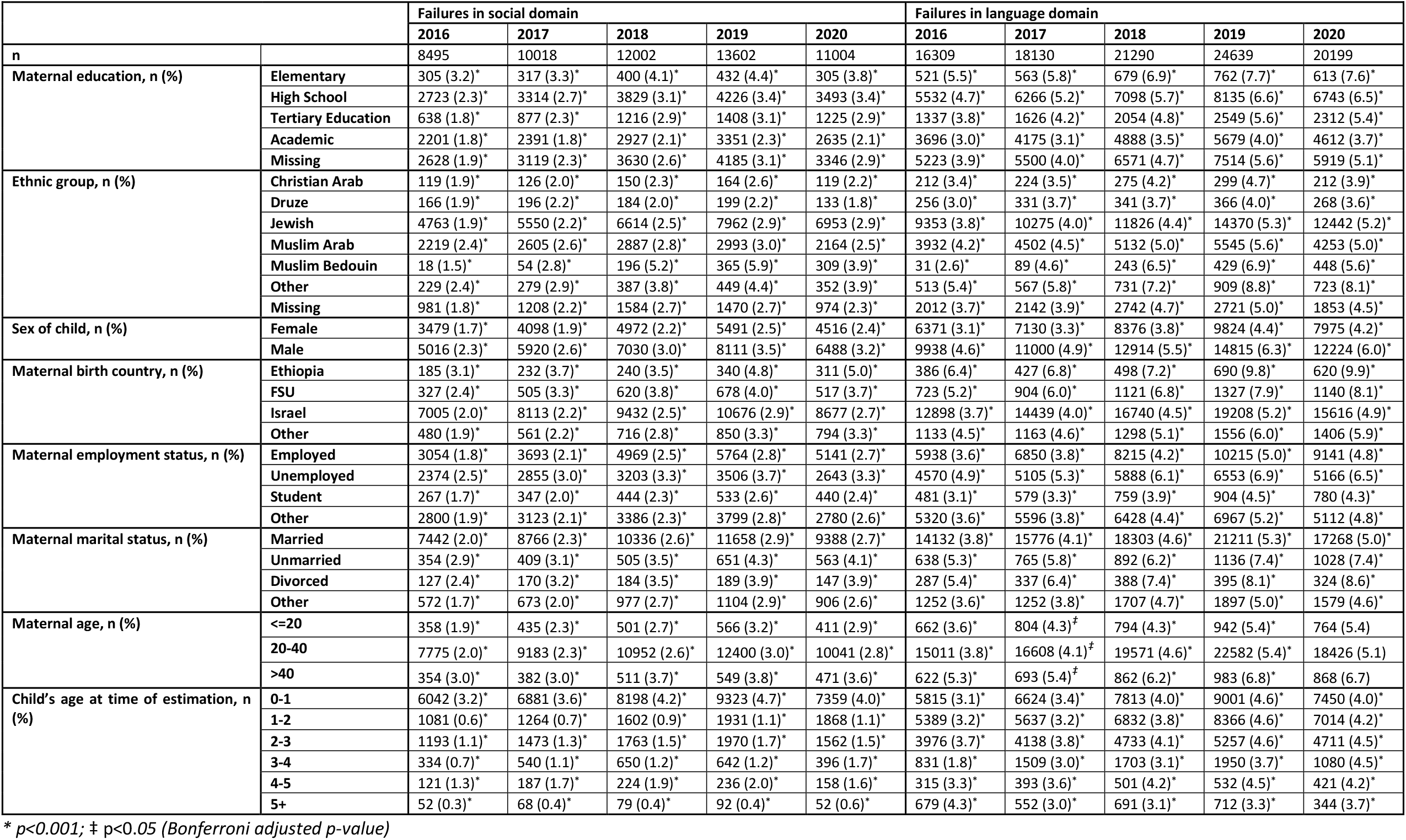
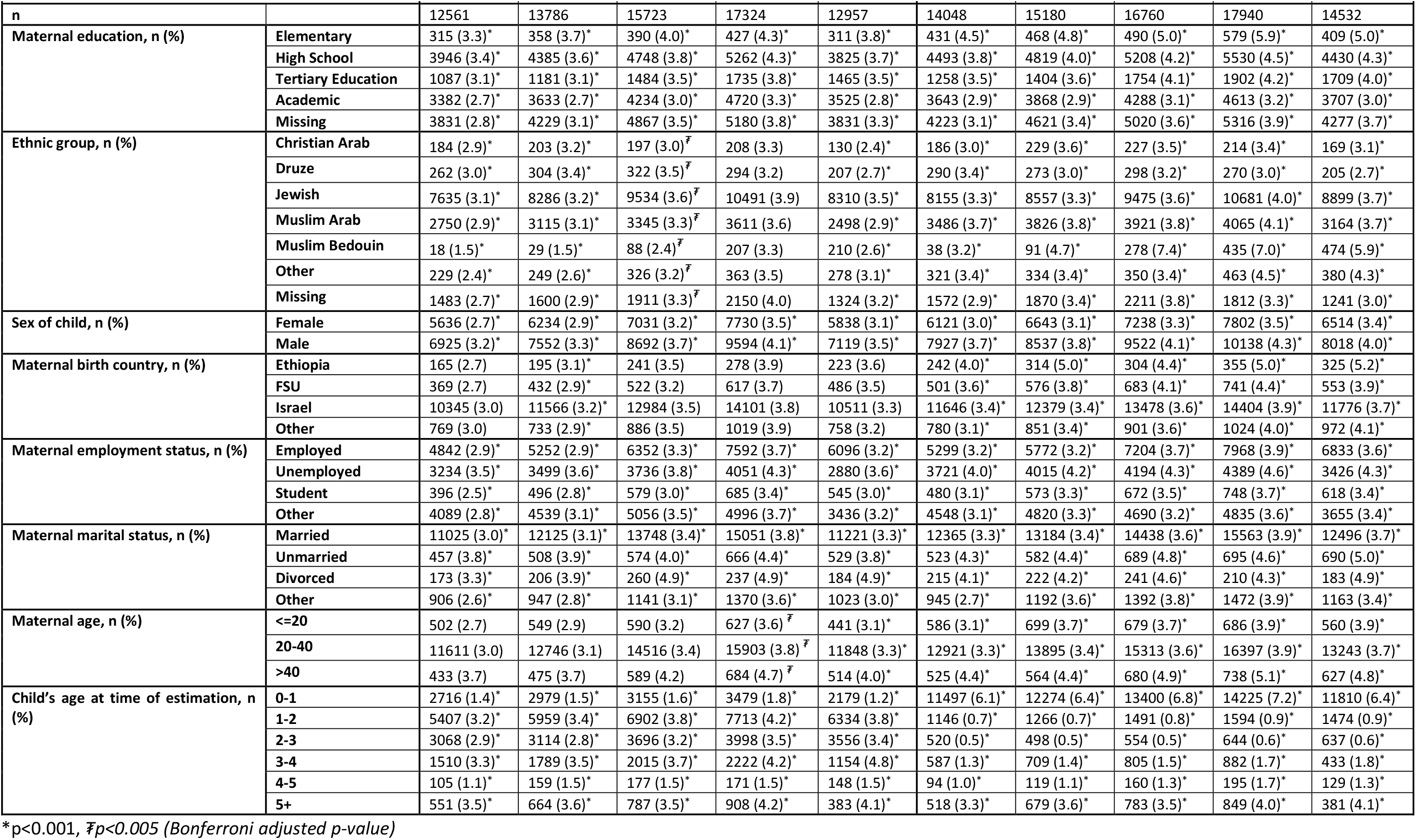
Study population characteristics by developmental domain attainment failure in four developmental domains: social, language, gross motor and fine motor

|  |  | Failures in social domain |  |  |  |  | Failures in language domain |  |  |  |  |
| --- | --- | --- | --- | --- | --- | --- | --- | --- | --- | --- | --- |
|  |  | 2016 | 2017 | 2018 | 2019 | 2020 | 2016 | 2017 | 2018 | 2019 | 2020 |
| **n** |  | 8495 | 10018 | 12002 | 13602 | 11004 | 16309 | 18130 | 21290 | 24639 | 20199 |
| **Maternal education, n (%)** | **Elementary** | 305 (3.2)* | 317 (3.3)* | 400 (4.1)* | 432 (4.4)* | 305 (3.8)* | 521 (5.5)* | 563 (5.8)* | 679 (6.9)* | 762 (7.7)* | 613 (7.6)* |
|  | **High School** | 2723 (2.3)* | 3314 (2.7)* | 3829 (3.1)* | 4226 (3.4)* | 3493 (3.4)* | 5532 (4.7)* | 6266 (5.2)* | 7098 (5.7)* | 8135 (6.6)* | 6743 (6.5)* |
|  | **Tertiary Education** | 638 (1.8)* | 877 (2.3)* | 1216 (2.9)* | 1408 (3.1)* | 1225 (2.9)* | 1337 (3.8)* | 1626 (4.2)* | 2054 (4.8)* | 2549 (5.6)* | 2312 (5.4)* |
|  | **Academic** | 2201 (1.8)* | 2391 (1.8)* | 2927 (2.1)* | 3351 (2.3)* | 2635 (2.1)* | 3696 (3.0)* | 4175 (3.1)* | 4888 (3.5)* | 5679 (4.0)* | 4612 (3.7)* |
|  | **Missing** | 2628 (1.9)* | 3119 (2.3)* | 3630 (2.6)* | 4185 (3.1)* | 3346 (2.9)* | 5223 (3.9)* | 5500 (4.0)* | 6571 (4.7)* | 7514 (5.6)* | 5919 (5.1)* |
| **Ethnic group, n (%)** | **Christian Arab** | 119 (1.9)* | 126 (2.0)* | 150 (2.3)* | 164 (2.6)* | 119 (2.2)* | 212 (3.4)* | 224 (3.5)* | 275 (4.2)* | 299 (4.7)* | 212 (3.9)* |
|  | **Druze** | 166 (1.9)* | 196 (2.2)* | 184 (2.0)* | 199 (2.2)* | 133 (1.8)* | 256 (3.0)* | 331 (3.7)* | 341 (3.7)* | 366 (4.0)* | 268 (3.6)* |
|  | **Jewish** | 4763 (1.9)* | 5550 (2.2)* | 6614 (2.5)* | 7962 (2.9)* | 6953 (2.9)* | 9353 (3.8)* | 10275 (4.0)* | 11826 (4.4)* | 14370 (5.3)* | 12442 (5.2)* |
|  | **Muslim Arab** | 2219 (2.4)* | 2605 (2.6)* | 2887 (2.8)* | 2993 (3.0)* | 2164 (2.5)* | 3932 (4.2)* | 4502 (4.5)* | 5132 (5.0)* | 5545 (5.6)* | 4253 (5.0)* |
|  | **Muslim Bedouin** | 18 (1.5)* | 54 (2.8)* | 196 (5.2)* | 365 (5.9)* | 309 (3.9)* | 31 (2.6)* | 89 (4.6)* | 243 (6.5)* | 429 (6.9)* | 448 (5.6)* |
|  | **Other** | 229 (2.4)* | 279 (2.9)* | 387 (3.8)* | 449 (4.4)* | 352 (3.9)* | 513 (5.4)* | 567 (5.8)* | 731 (7.2)* | 909 (8.8)* | 723 (8.1)* |
|  | **Missing** | 981 (1.8)* | 1208 (2.2)* | 1584 (2.7)* | 1470 (2.7)* | 974 (2.3)* | 2012 (3.7)* | 2142 (3.9)* | 2742 (4.7)* | 2721 (5.0)* | 1853 (4.5)* |
| **Sex of child, n (%)** | **Female** | 3479 (1.7)* | 4098 (1.9)* | 4972 (2.2)* | 5491 (2.5)* | 4516 (2.4)* | 6371 (3.1)* | 7130 (3.3)* | 8376 (3.8)* | 9824 (4.4)* | 7975 (4.2)* |
|  | **Male** | 5016 (2.3)* | 5920 (2.6)* | 7030 (3.0)* | 8111 (3.5)* | 6488 (3.2)* | 9938 (4.6)* | 11000 (4.9)* | 12914 (5.5)* | 14815 (6.3)* | 12224 (6.0)* |
| **Maternal birth country, n (%)** | **Ethiopia** | 185 (3.1)* | 232 (3.7)* | 240 (3.5)* | 340 (4.8)* | 311 (5.0)* | 386 (6.4)* | 427 (6.8)* | 498 (7.2)* | 690 (9.8)* | 620 (9.9)* |
|  | **FSU** | 327 (2.4)* | 505 (3.3)* | 620 (3.8)* | 678 (4.0)* | 517 (3.7)* | 723 (5.2)* | 904 (6.0)* | 1121 (6.8)* | 1327 (7.9)* | 1140 (8.1)* |
|  | **Israel** | 7005 (2.0)* | 8113 (2.2)* | 9432 (2.5)* | 10676 (2.9)* | 8677 (2.7)* | 12898 (3.7)* | 14439 (4.0)* | 16740 (4.5)* | 19208 (5.2)* | 15616 (4.9)* |
|  | **Other** | 480 (1.9)* | 561 (2.2)* | 716 (2.8)* | 850 (3.3)* | 794 (3.3)* | 1133 (4.5)* | 1163 (4.6)* | 1298 (5.1)* | 1556 (6.0)* | 1406 (5.9)* |
| **Maternal employment status, n (%)** | **Employed** | 3054 (1.8)* | 3693 (2.1)* | 4969 (2.5)* | 5764 (2.8)* | 5141 (2.7)* | 5938 (3.6)* | 6850 (3.8)* | 8215 (4.2)* | 10215 (5.0)* | 9141 (4.8)* |
|  | **Unemployed** | 2374 (2.5)* | 2855 (3.0)* | 3203 (3.3)* | 3506 (3.7)* | 2643 (3.3)* | 4570 (4.9)* | 5105 (5.3)* | 5888 (6.1)* | 6553 (6.9)* | 5166 (6.5)* |
|  | **Student** | 267 (1.7)* | 347 (2.0)* | 444 (2.3)* | 533 (2.6)* | 440 (2.4)* | 481 (3.1)* | 579 (3.3)* | 759 (3.9)* | 904 (4.5)* | 780 (4.3)* |
|  | **Other** | 2800 (1.9)* | 3123 (2.1)* | 3386 (2.3)* | 3799 (2.8)* | 2780 (2.6)* | 5320 (3.6)* | 5596 (3.8)* | 6428 (4.4)* | 6967 (5.2)* | 5112 (4.8)* |
| **Maternal marital status, n (%)** | **Married** | 7442 (2.0)* | 8766 (2.3)* | 10336 (2.6)* | 11658 (2.9)* | 9388 (2.7)* | 14132 (3.8)* | 15776 (4.1)* | 18303 (4.6)* | 21211 (5.3)* | 17268 (5.0)* |
|  | **Unmarried** | 354 (2.9)* | 409 (3.1)* | 505 (3.5)* | 651 (4.3)* | 563 (4.1)* | 638 (5.3)* | 765 (5.8)* | 892 (6.2)* | 1136 (7.4)* | 1028 (7.4)* |
|  | **Divorced** | 127 (2.4)* | 170 (3.2)* | 184 (3.5)* | 189 (3.9)* | 147 (3.9)* | 287 (5.4)* | 337 (6.4)* | 388 (7.4)* | 395 (8.1)* | 324 (8.6)* |
|  | **Other** | 572 (1.7)* | 673 (2.0)* | 977 (2.7)* | 1104 (2.9)* | 906 (2.6)* | 1252 (3.6)* | 1252 (3.8)* | 1707 (4.7)* | 1897 (5.0)* | 1579 (4.6)* |
| **Maternal age, n (%)** | **<=20** | 358 (1.9)* | 435 (2.3)* | 501 (2.7)* | 566 (3.2)* | 411 (2.9)* | 662 (3.6)* | 804 (4.3)<sup>‡</sup> | 794 (4.3)* | 942 (5.4)* | 764 (5.4) |
|  | **20-40** | 7775 (2.0)* | 9183 (2.3)* | 10952 (2.6)* | 12400 (3.0)* | 10041 (2.8)* | 15011 (3.8)* | 16608 (4.1)<sup>‡</sup> | 19571 (4.6)* | 22582 (5.4)* | 18426 (5.1) |
|  | **>40** | 354 (3.0)* | 382 (3.0)* | 511 (3.7)* | 549 (3.8)* | 471 (3.6)* | 622 (5.3)* | 693 (5.4)<sup>‡</sup> | 862 (6.2)* | 983 (6.8)* | 868 (6.7) |
| **Child's age at time of estimation, n (%)** | **0-1** | 6042 (3.2)* | 6881 (3.6)* | 8198 (4.2)* | 9323 (4.7)* | 7359 (4.0)* | 5815 (3.1)* | 6624 (3.4)* | 7813 (4.0)* | 9001 (4.6)* | 7450 (4.0)* |
|  | **1-2** | 1081 (0.6)* | 1264 (0.7)* | 1602 (0.9)* | 1931 (1.1)* | 1868 (1.1)* | 5389 (3.2)* | 5637 (3.2)* | 6832 (3.8)* | 8366 (4.6)* | 7014 (4.2)* |
|  | **2-3** | 1193 (1.1)* | 1473 (1.3)* | 1763 (1.5)* | 1970 (1.7)* | 1562 (1.5)* | 3976 (3.7)* | 4138 (3.8)* | 4733 (4.1)* | 5257 (4.6)* | 4711 (4.5)* |
|  | **3-4** | 334 (0.7)* | 540 (1.1)* | 650 (1.2)* | 642 (1.2)* | 396 (1.7)* | 831 (1.8)* | 1509 (3.0)* | 1703 (3.1)* | 1950 (3.7)* | 1080 (4.5)* |
|  | **4-5** | 121 (1.3)* | 187 (1.7)* | 224 (1.9)* | 236 (2.0)* | 158 (1.6)* | 315 (3.3)* | 393 (3.6)* | 501 (4.2)* | 532 (4.5)* | 421 (4.2)* |
|  | **5+** | 52 (0.3)* | 68 (0.4)* | 79 (0.4)* | 92 (0.4)* | 52 (0.6)* | 679 (4.3)* | 552 (3.0)* | 691 (3.1)* | 712 (3.3)* | 344 (3.7)* |

\* $p < 0.001$ ; ‡ $p < 0.05$ (Bonferroni adjusted $p$ -value)

Table 1.c. Study population characteristics by developmental domain attainment failure in four developmental domains: social, language, gross motor and fine motor (continued)
|  |  | Failures in gross motor domain |  |  |  |  | Failures in fine motor domain |  |  |  |  |
| --- | --- | --- | --- | --- | --- | --- | --- | --- | --- | --- | --- |
|  |  | 2016 | 2017 | 2018 | 2019 | 2020 | 2016 | 2017 | 2018 | 2019 | 2020 |
| n |  | 12561 | 13786 | 15723 | 17324 | 12957 | 14048 | 15180 | 16760 | 17940 | 14532 |
| Maternal education, n (%) | Elementary | 315 (3.3)* | 358 (3.7)* | 390 (4.0)* | 427 (4.3)* | 311 (3.8)* | 431 (4.5)* | 468 (4.8)* | 490 (5.0)* | 579 (5.9)* | 409 (5.0)* |
|  | High School | 3946 (3.4)* | 4385 (3.6)* | 4748 (3.8)* | 5262 (4.3)* | 3825 (3.7)* | 4493 (3.8)* | 4819 (4.0)* | 5208 (4.2)* | 5530 (4.5)* | 4430 (4.3)* |
|  | Tertiary Education | 1087 (3.1)* | 1181 (3.1)* | 1484 (3.5)* | 1735 (3.8)* | 1465 (3.5)* | 1258 (3.5)* | 1404 (3.6)* | 1754 (4.1)* | 1902 (4.2)* | 1709 (4.0)* |
|  | Academic | 3382 (2.7)* | 3633 (2.7)* | 4234 (3.0)* | 4720 (3.3)* | 3525 (2.8)* | 3643 (2.9)* | 3868 (2.9)* | 4288 (3.1)* | 4613 (3.2)* | 3707 (3.0)* |
|  | Missing | 3831 (2.8)* | 4229 (3.1)* | 4867 (3.5)* | 5180 (3.8)* | 3831 (3.3)* | 4223 (3.1)* | 4621 (3.4)* | 5020 (3.6)* | 5316 (3.9)* | 4277 (3.7)* |
| Ethnic group, n (%) | Christian Arab | 184 (2.9)* | 203 (3.2)* | 197 (3.0)<sup>‡</sup> | 208 (3.3) | 130 (2.4)* | 186 (3.0)* | 229 (3.6)* | 227 (3.5)* | 214 (3.4)* | 169 (3.1)* |
|  | Druze | 262 (3.0)* | 304 (3.4)* | 322 (3.5)<sup>‡</sup> | 294 (3.2) | 207 (2.7)* | 290 (3.4)* | 273 (3.0)* | 298 (3.2)* | 270 (3.0)* | 205 (2.7)* |
|  | Jewish | 7635 (3.1)* | 8286 (3.2)* | 9534 (3.6)<sup>‡</sup> | 10491 (3.9) | 8310 (3.5)* | 8155 (3.3)* | 8557 (3.3)* | 9475 (3.6)* | 10681 (4.0)* | 8899 (3.7)* |
|  | Muslim Arab | 2750 (2.9)* | 3115 (3.1)* | 3345 (3.3)<sup>‡</sup> | 3611 (3.6) | 2498 (2.9)* | 3486 (3.7)* | 3826 (3.8)* | 3921 (3.8)* | 4065 (4.1)* | 3164 (3.7)* |
|  | Muslim Bedouin | 18 (1.5)* | 29 (1.5)* | 88 (2.4)<sup>‡</sup> | 207 (3.3) | 210 (2.6)* | 38 (3.2)* | 91 (4.7)* | 278 (7.4)* | 435 (7.0)* | 474 (5.9)* |
|  | Other | 229 (2.4)* | 249 (2.6)* | 326 (3.2)<sup>‡</sup> | 363 (3.5) | 278 (3.1)* | 321 (3.4)* | 334 (3.4)* | 350 (3.4)* | 463 (4.5)* | 380 (4.3)* |
|  | Missing | 1483 (2.7)* | 1600 (2.9)* | 1911 (3.3)<sup>‡</sup> | 2150 (4.0) | 1324 (3.2)* | 1572 (2.9)* | 1870 (3.4)* | 2211 (3.8)* | 1812 (3.3)* | 1241 (3.0)* |
| Sex of child, n (%) | Female | 5636 (2.7)* | 6234 (2.9)* | 7031 (3.2)* | 7730 (3.5)* | 5838 (3.1)* | 6121 (3.0)* | 6643 (3.1)* | 7238 (3.3)* | 7802 (3.5)* | 6514 (3.4)* |
|  | Male | 6925 (3.2)* | 7552 (3.3)* | 8692 (3.7)* | 9594 (4.1)* | 7119 (3.5)* | 7927 (3.7)* | 8537 (3.8)* | 9522 (4.1)* | 10138 (4.3)* | 8018 (4.0)* |
| Maternal birth country, n (%) | Ethiopia | 165 (2.7) | 195 (3.1)* | 241 (3.5) | 278 (3.9) | 223 (3.6) | 242 (4.0)* | 314 (5.0)* | 304 (4.4)* | 355 (5.0)* | 325 (5.2)* |
|  | FSU | 369 (2.7) | 432 (2.9)* | 522 (3.2) | 617 (3.7) | 486 (3.5) | 501 (3.6)* | 576 (3.8)* | 683 (4.1)* | 741 (4.4)* | 553 (3.9)* |
|  | Israel | 10345 (3.0) | 11566 (3.2)* | 12984 (3.5) | 14101 (3.8) | 10511 (3.3) | 11646 (3.4)* | 12379 (3.4)* | 13478 (3.6)* | 14404 (3.9)* | 11776 (3.7)* |
|  | Other | 769 (3.0) | 733 (2.9)* | 886 (3.5) | 1019 (3.9) | 758 (3.2) | 780 (3.1)* | 851 (3.4)* | 901 (3.6)* | 1024 (4.0)* | 972 (4.1)* |
| Maternal employment status, n (%) | Employed | 4842 (2.9)* | 5252 (2.9)* | 6352 (3.3)* | 7592 (3.7)* | 6096 (3.2)* | 5299 (3.2)* | 5772 (3.2)* | 7204 (3.7)* | 7968 (3.9)* | 6833 (3.6)* |
|  | Unemployed | 3234 (3.5)* | 3499 (3.6)* | 3736 (3.8)* | 4051 (4.3)* | 2880 (3.6)* | 3721 (4.0)* | 4015 (4.2)* | 4194 (4.3)* | 4389 (4.6)* | 3426 (4.3)* |
|  | Student | 396 (2.5)* | 496 (2.8)* | 579 (3.0)* | 685 (3.4)* | 545 (3.0)* | 480 (3.1)* | 573 (3.3)* | 672 (3.5)* | 748 (3.7)* | 618 (3.4)* |
|  | Other | 4089 (2.8)* | 4539 (3.1)* | 5056 (3.5)* | 4996 (3.7)* | 3436 (3.2)* | 4548 (3.1)* | 4820 (3.3)* | 4690 (3.2)* | 4835 (3.6)* | 3655 (3.4)* |
| Maternal marital status, n (%) | Married | 11025 (3.0)* | 12125 (3.1)* | 13748 (3.4)* | 15051 (3.8)* | 11221 (3.3)* | 12365 (3.3)* | 13184 (3.4)* | 14438 (3.6)* | 15563 (3.9)* | 12496 (3.7)* |
|  | Unmarried | 457 (3.8)* | 508 (3.9)* | 574 (4.0)* | 666 (4.4)* | 529 (3.8)* | 523 (4.3)* | 582 (4.4)* | 689 (4.8)* | 695 (4.6)* | 690 (5.0)* |
|  | Divorced | 173 (3.3)* | 206 (3.9)* | 260 (4.9)* | 237 (4.9)* | 184 (4.9)* | 215 (4.1)* | 222 (4.2)* | 241 (4.6)* | 210 (4.3)* | 183 (4.9)* |
|  | Other | 906 (2.6)* | 947 (2.8)* | 1141 (3.1)* | 1370 (3.6)* | 1023 (3.0)* | 945 (2.7)* | 1192 (3.6)* | 1392 (3.8)* | 1472 (3.9)* | 1163 (3.4)* |
| Maternal age, n (%) | <=20 | 502 (2.7) | 549 (2.9) | 590 (3.2) | 627 (3.6)<sup>‡</sup> | 441 (3.1)* | 586 (3.1)* | 699 (3.7)* | 679 (3.7)* | 686 (3.9)* | 560 (3.9)* |
|  | 20-40 | 11611 (3.0) | 12746 (3.1) | 14516 (3.4) | 15903 (3.8)<sup>‡</sup> | 11848 (3.3)* | 12921 (3.3)* | 13895 (3.4)* | 15313 (3.6)* | 16397 (3.9)* | 13243 (3.7)* |
|  | >40 | 433 (3.7) | 475 (3.7) | 589 (4.2) | 684 (4.7)<sup>‡</sup> | 514 (4.0)* | 525 (4.4)* | 564 (4.4)* | 680 (4.9)* | 738 (5.1)* | 627 (4.8)* |
| Child's age at time of estimation, n (%) | 0-1 | 2716 (1.4)* | 2979 (1.5)* | 3155 (1.6)* | 3479 (1.8)* | 2179 (1.2)* | 11497 (6.1)* | 12274 (6.4)* | 13400 (6.8)* | 14225 (7.2)* | 11810 (6.4)* |
|  | 1-2 | 5407 (3.2)* | 5959 (3.4)* | 6902 (3.8)* | 7713 (4.2)* | 6334 (3.8)* | 1146 (0.7)* | 1266 (0.7)* | 1491 (0.8)* | 1594 (0.9)* | 1474 (0.9)* |
|  | 2-3 | 3068 (2.9)* | 3114 (2.8)* | 3696 (3.2)* | 3998 (3.5)* | 3556 (3.4)* | 520 (0.5)* | 498 (0.5)* | 554 (0.5)* | 644 (0.6)* | 637 (0.6)* |
|  | 3-4 | 1510 (3.3)* | 1789 (3.5)* | 2015 (3.7)* | 2222 (4.2)* | 1154 (4.8)* | 587 (1.3)* | 709 (1.4)* | 805 (1.5)* | 882 (1.7)* | 433 (1.8)* |
|  | 4-5 | 105 (1.1)* | 159 (1.5)* | 177 (1.5)* | 171 (1.5)* | 148 (1.5)* | 94 (1.0)* | 119 (1.1)* | 160 (1.3)* | 195 (1.7)* | 129 (1.3)* |
|  | 5+ | 551 (3.5)* | 664 (3.6)* | 787 (3.5)* | 908 (4.2)* | 383 (4.1)* | 518 (3.3)* | 679 (3.6)* | 783 (3.5)* | 849 (4.0)* | 381 (4.1)* |

\*p<0.001, <sup>‡</sup>p<0.005 (Bonferroni adjusted p-value)

### Statistical analysis

We report the annual percentage of children with the outcome overall, and stratified by sociodemographic factors, along with 95% confidence intervals for proportions. This data is presented nationally as well as in geographical sub-division.

Descriptive statistics on baseline population characteristics were generated with percentage. Comparisons between sociodemographic sub-groups were analyzed with χ2 tests adjusted for multiple comparisons via Bonferroni correction. The statistical significance of the trends in failure incidence rates was calculated using the Cochran–Armitage trend test.

We used multivariable logistic regression to estimate the impact of each explanatory variable on the odds ratio (OR) of failure to attain developmental milestones, while controlling for confounding variables. We repeated this analysis using failure in each of the developmental domains as the response variable. We used variance inflation factor (VIF) to assess potential multicollinearity between the explanatory variables. The analysis was performed using Python programming language version 3.6 (Python Software Foundation).

## Results

1,002,700 children with 4,441,689 developmental visits were included. Out of these visits, 535 were excluded due to lack of developmental data, missing age or duplicate records.

The cohort’s main characteristics are presented in Table 1(a,b,c). This cohort included approximately 70% of the children born in Israel within the study period, and is mostly representative of the entire national population, with a slight overrepresentation of the Arab sector (76%-80% of the Israeli-Arab children are included vs. 59%-68% of the Israeli-Jewish children) [20].

### General trends

Over the years 2016-2019, there was an increase in the rate of failure to attain milestones in all four developmental domains (p<0.001), as shown in Figure 1. The milestone attainment failure rate was ranging from 3.1% [2.92, 3.02] in 2016 to 3.8% [3.74, 3.86] in 2019 in the gross motor domain, from 3.3% [3.27, 3.38] in 2016 to 3.9% [3.88, 3.99] in 2019 in the fine motor domain, and from 2% [1.97, 2.05] in 2016 to 3% [2.93, 3.03] in 2019 in the social domain. The highest failure rate was observed in language milestones, with a rate of 3.9% [3.8, 3.92] in 2016 and 5.4% [5.34, 5.47] in 2019. In 2020 there was a slight decrease in failure rates, however, this may be a consequence of changes in the surveillance policy and availability between March and October 2020 due to constraints of the COVID-19 pandemic. Parental concern progressively decreased over the years, from 3.9% [3.89, 4.01] in 2016 to 3.3% [3.23, 3.34] in 2020.

**Figure 1.**
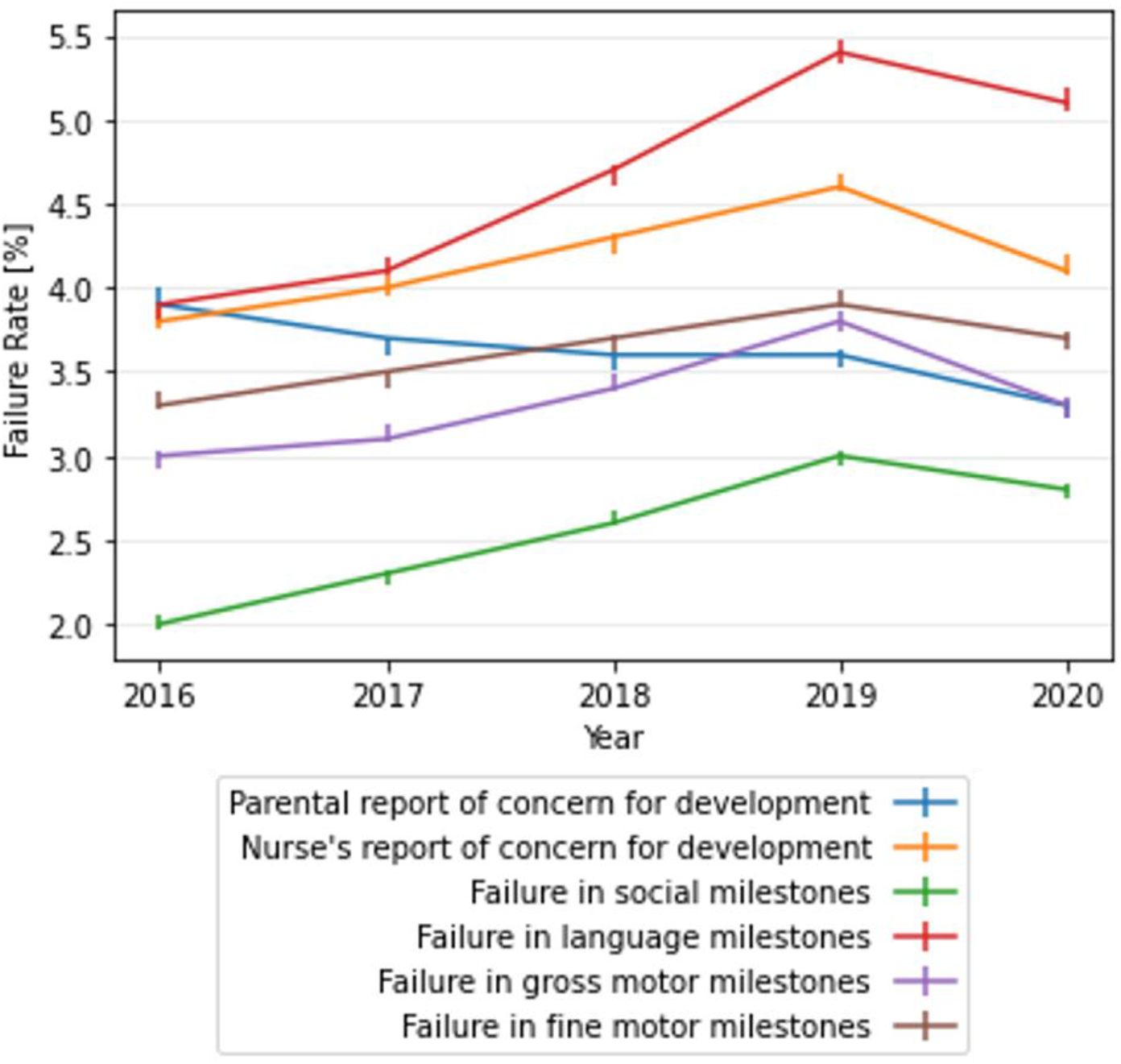
Temporal trends in child development domains and in parental and nurse’s report of concern for child development

### Sociodemographic correlates of child development

#### Maternal education

Figure 2 shows the temporal trends of developmental outcomes stratified by the mother’s level of education. It is apparent that the failure rate in all developmental domains is lower as the mother’s level of education is higher. This trend is more prominent in the language domain (Figure 2(d)) than in the gross motor domain (Figure 2(e)). The same trend was observed in the nurse’s report of developmental concern (Figure 2(b)). In the language, social and fine motor domains, the rate of failure to attain milestones is increasing more rapidly for children with lower maternal education, widening the gap between the groups (Figure 2(c,d,f)). The language milestone attainment failure rate in children whose mothers had elementary education increased from 5.5% in 2016 to 7.7% and 7.6% in 2019 and 2020, respectively. In children whose mothers had academic education, the milestone attainment failure rate increased from 3% in 2016 to 4% and 3.7% in 2019 and 2020, respectively (p<0.001). In milestones in the fine motor and social domains the attainment failure rates in 2016 were 3.2% and 4.5%, respectively, in children whose mothers had elementary education, and 1.8% and 2.9% respectively, in children whose mothers had academic education (p<0.001). In 2019 and 2020 the attainment failure rates for children whose mothers had elementary education were 4.4% and 3.8% respectively, in the social domain, and 5.9% and 5%, respectively, in the fine motor domain. The attainment failure rates for children whose mothers had academic education in 2019 and 2020 were 2.3% and 2.1%, respectively, in the social domain, and 3.2% and 3%, respectively, in the fine motor domain (p<0.001).

**Figure 2.**
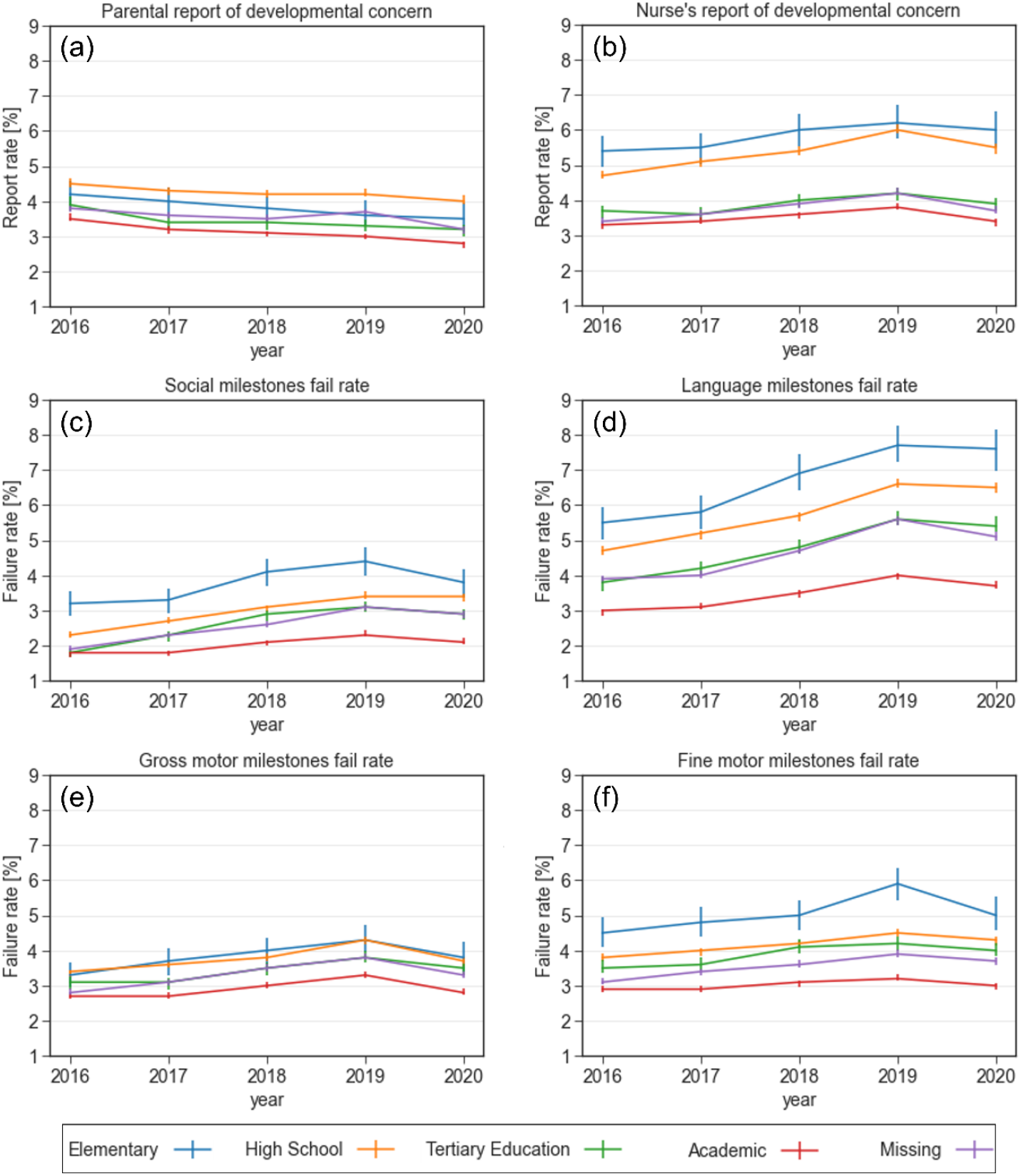
Reports of concern and failure rates in various developmental domains stratified by maternal education level between 2016-2020. (a) Parental report of developmental concern (b) Nurse’s report of developmental concern (c) Failure rate in the social domain (d) Failure rate in the language domain (e) Failure rate in the gross motor domain (f) Failure rate in the fine motor domain

#### Ethnic group

Figure 3 shows the trend of developmental outcomes stratified by the different ethnic and religious groups, including Jews, Muslim Arabs, Christian Arabs, Druze, and Bedouin. The Bedouin children had higher failure rates in the social (3.9%, p<0.001), language (5.6%, p<0.001), and fine motor (5.9%, p<0.001) domains (Figure 3(c,d,f)). In contrast, the level of concern among this group, by both the parents and the attending nurse was the lowest (Figure 3(b)), reflecting higher unawareness to potential developmental delays. Interestingly, the highest rates of milestone attainment in these domains were observed in another minority group, the Druze, while the rate of concern by the nurses was highest for this group.

**Figure 3.**
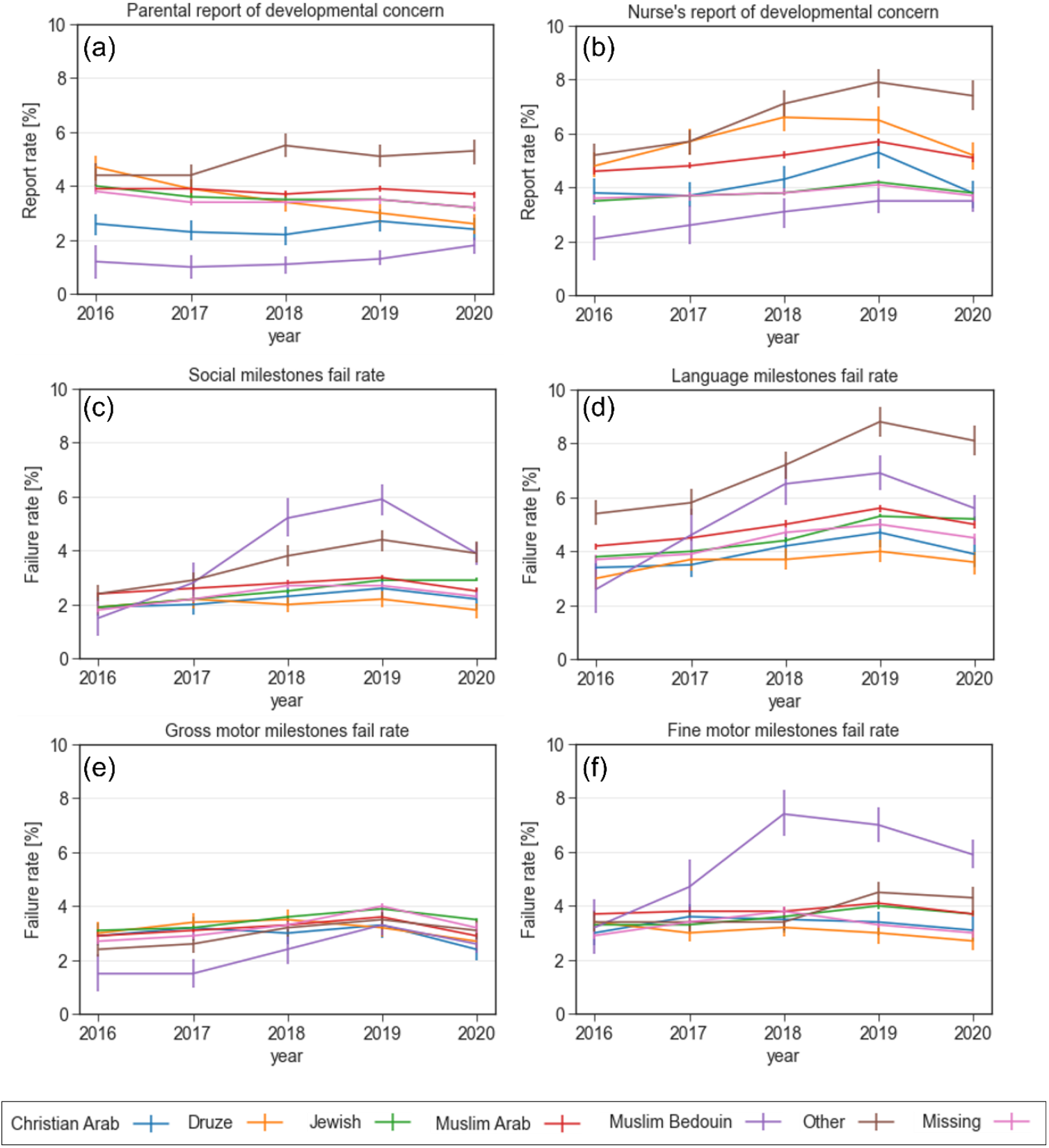
Reports of concern and failure rates in various developmental domains stratified by ethnic and religious groups between 2016-2020. (a) Parental report of developmental concern (b) Nurse’s report of developmental concern (c) Failure rate in the social domain (d) Failure rate in the language domain (e) Failure rate in the gross motor domain (f) Failure rate in the fine motor domain

#### Maternal birth country

Figure 4 shows the trend of developmental outcomes stratified by maternal birth country. Failure rates in attaining social and language milestones were higher for children whose mothers immigrated to Israel (mostly from the Former Soviet Union (FSU) or Ethiopia), while the failure rate in attaining fine motor milestones was higher for children of Ethiopia-born mothers (5.2% in 2020, p<0.001). Parental concern regarding their child’s development slightly increased over the years among mothers born in the FSU (p<0.001), and their rate was higher compared to mothers born in Israel or other countries (5% vs. 3%-3.6%, respectively, p<0.001). In nurse’s report of developmental concern, the rates of children to FSU- or Ethiopia-born mothers were substantially higher compared to children of Israel-born mothers (7%-7.2% vs. 4.0%, respectively. p<0.001).

**Figure 4.**
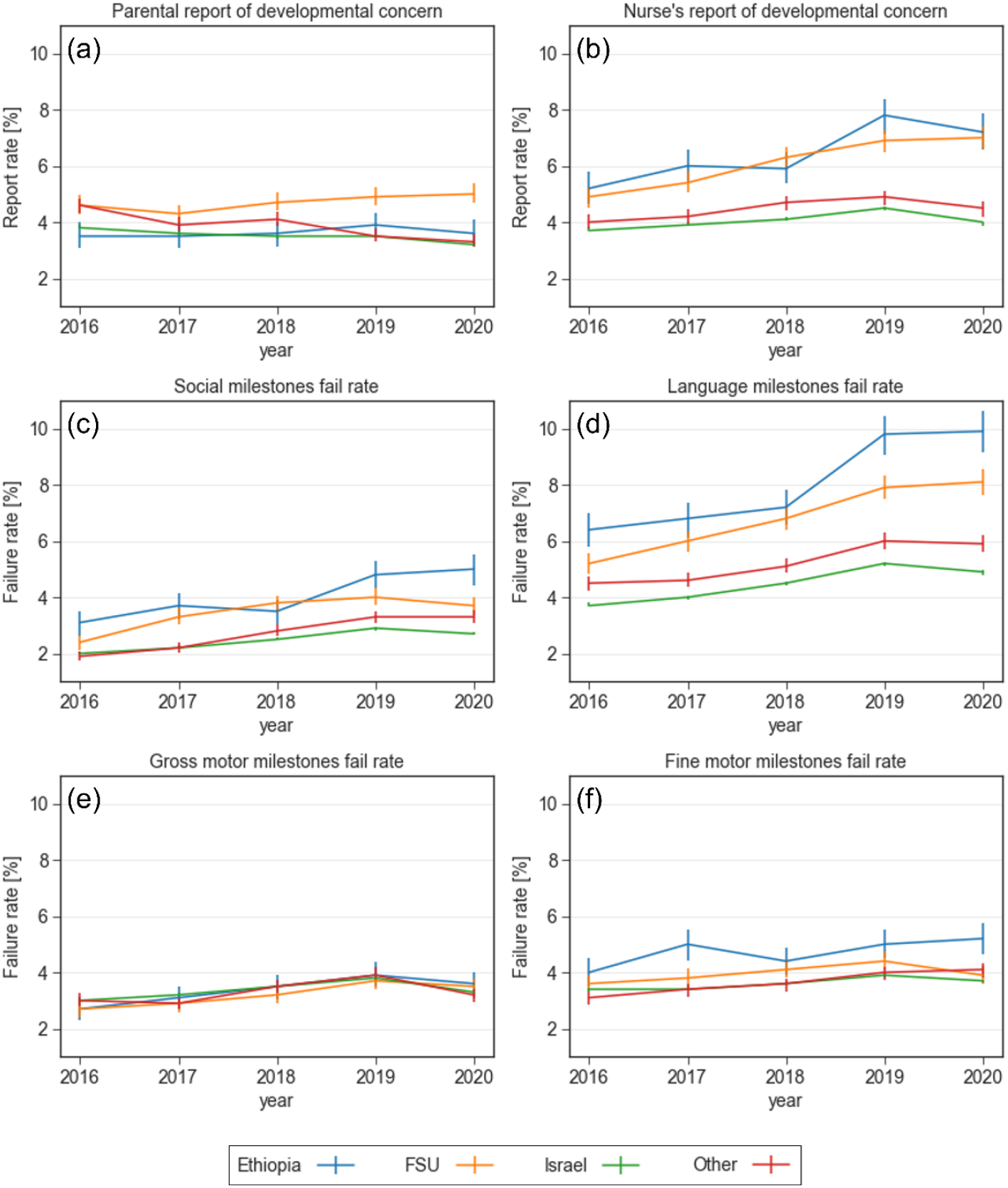
Reports of concern and failure rates in various developmental domains stratified by maternal country of birth between 2016-2020. (a) Parental report of developmental concern (b) Nurse’s report of developmental concern (c) Failure rate in the social domain (d) Failure rate in the language domain (e) Failure rate in the gross motor domain (f) Failure rate in the fine motor domain

#### Maternal employment status

Figure 5 describes the association between maternal employment status and the developmental outcomes. In all four domains, failure to attain milestones was more prevalent in children to unemployed mothers, compared with children to employed or student mothers. This difference was most prominent in the language milestones, where the failure rates were 6.9% and 6.5% (P<0.001) in 2019 and 2020, respectively, for children to unemployed mothers, compared to children to employed mothers with failure rates of 5% in 2019 and 4.8% in 2020 (p<0.001). Similarly, unemployed mothers reported concern regarding their child’s development more frequently than employed mothers or students.

**Figure 5.**
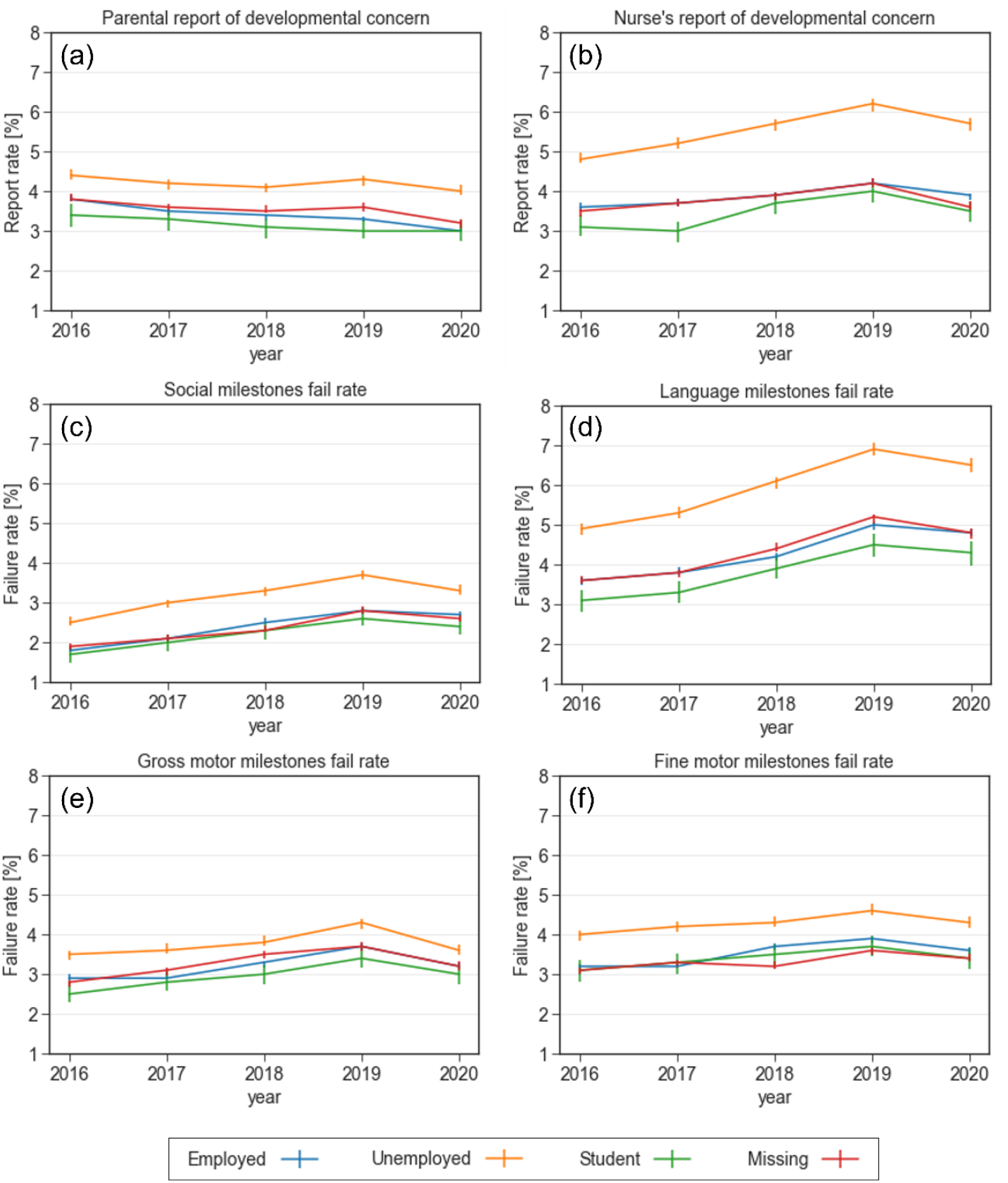
Reports of concern and failure rates in various developmental domains stratified by maternal employment status between 2016-2020. (a) Parental report of developmental concern (b) Nurse’s report of developmental concern (c) Failure rate in the social domain (d) Failure rate in the language domain (e) Failure rate in the gross motor domain (f) Failure rate in the fine motor domain

#### Maternal marital status

Figure 6 describes the association between maternal marital status and developmental outcomes. Married women were less likely to have concerns regarding their child’s development (3.2%, p<0.001 in 2020), compared to unmarried (4.5%, p<0.001 in 2020) or divorced (5.6%, p<0.001 in 2020) women (Figure 6 (a)). Similar associations were observed in nurse’s report regarding developmental concern (Figure 6(b)) and in objective failure in developmental milestones across all four domains according to THIS scale (Figure 6(c)-(f)).

**Figure 6.**
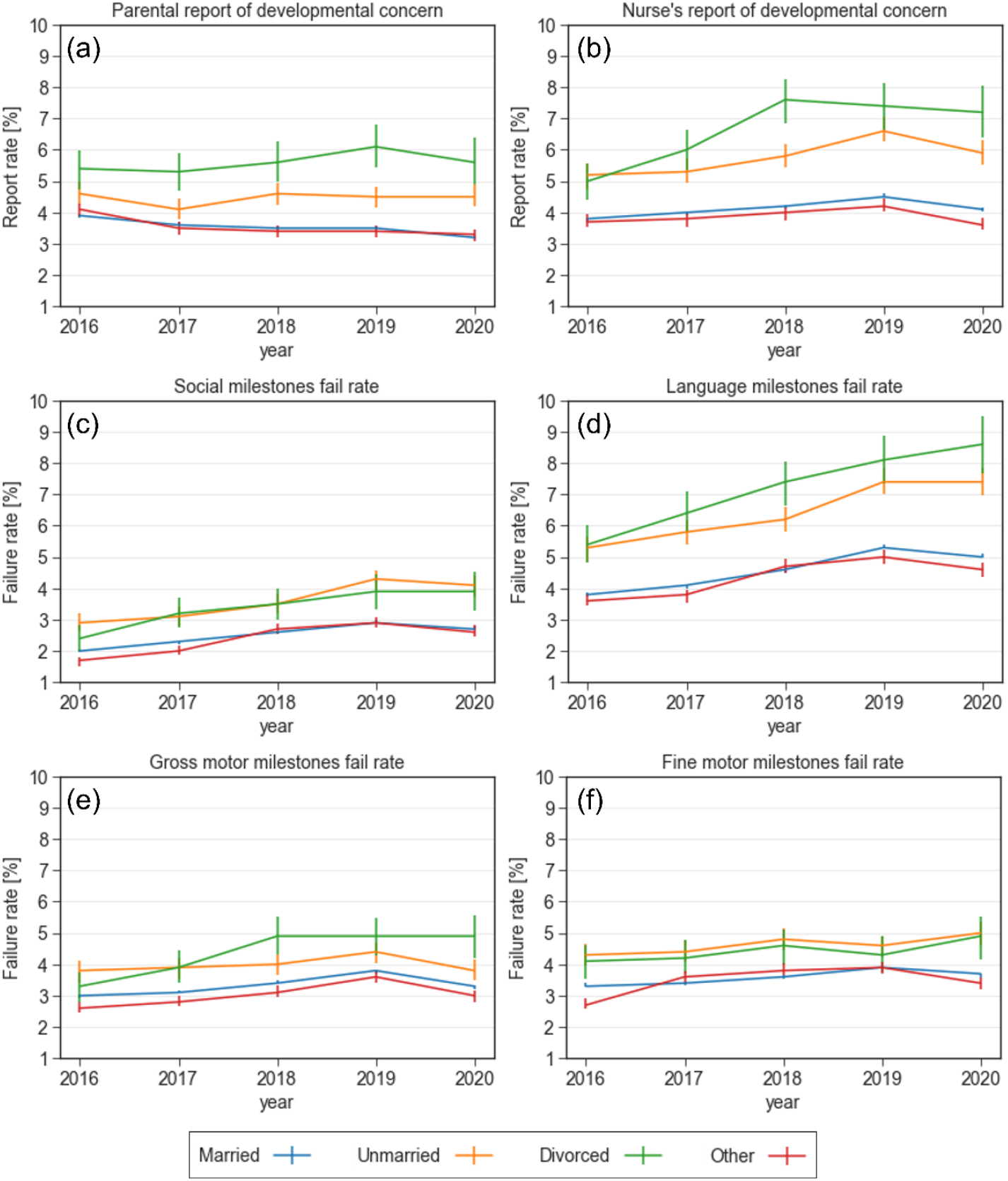
Reports of concern and failure rates in various developmental domains stratified by maternal marital status between 2016-2020. (a) Parental report of developmental concern (b) Nurse’s report of developmental concern (c) Failure rate in the social domain (d) Failure rate in the language domain (e) Failure rate in the gross motor domain (f) Failure rate in the fine motor domain

#### Sex of child

Failure rates were higher for males, compared to females, in all the developmental domains (Supplementary Figure 1). The gap between sex groups was largest in the language domain (for example, in 2020, 4.2% of assessed females failed in attaining language milestones, compared to 6% of assessed males, P<0.001).

#### Maternal age

Reports of concern and failure rates in various developmental domains stratified by maternal age between 2016-2020 are provided in supplementary Figure 2. Women who gave birth over the age of 40 report more concern regarding their child’s development, compared to women who gave birth between 20 and 40 years or those who gave birth before the age of 20 (p<0.001). Nurses report greater developmental concern in children whose mothers gave birth over the age of 40 (Supplementary Figure 2(b)). Similar associations were observed in objective failure in developmental milestones across all four domains (Supplementary Figure 2(c)-(f)).

Additional sub-analysis by geographical districts is available online (in Hebrew) [21].

### Contribution of sociodemographic variables to developmental milestone attainment

Mutually adjusted odds ratios for failure in any milestone, and for domain-specific failure are presented in Figure 7 and supplementary figures 3-6, respectively.

**Figure 7.**
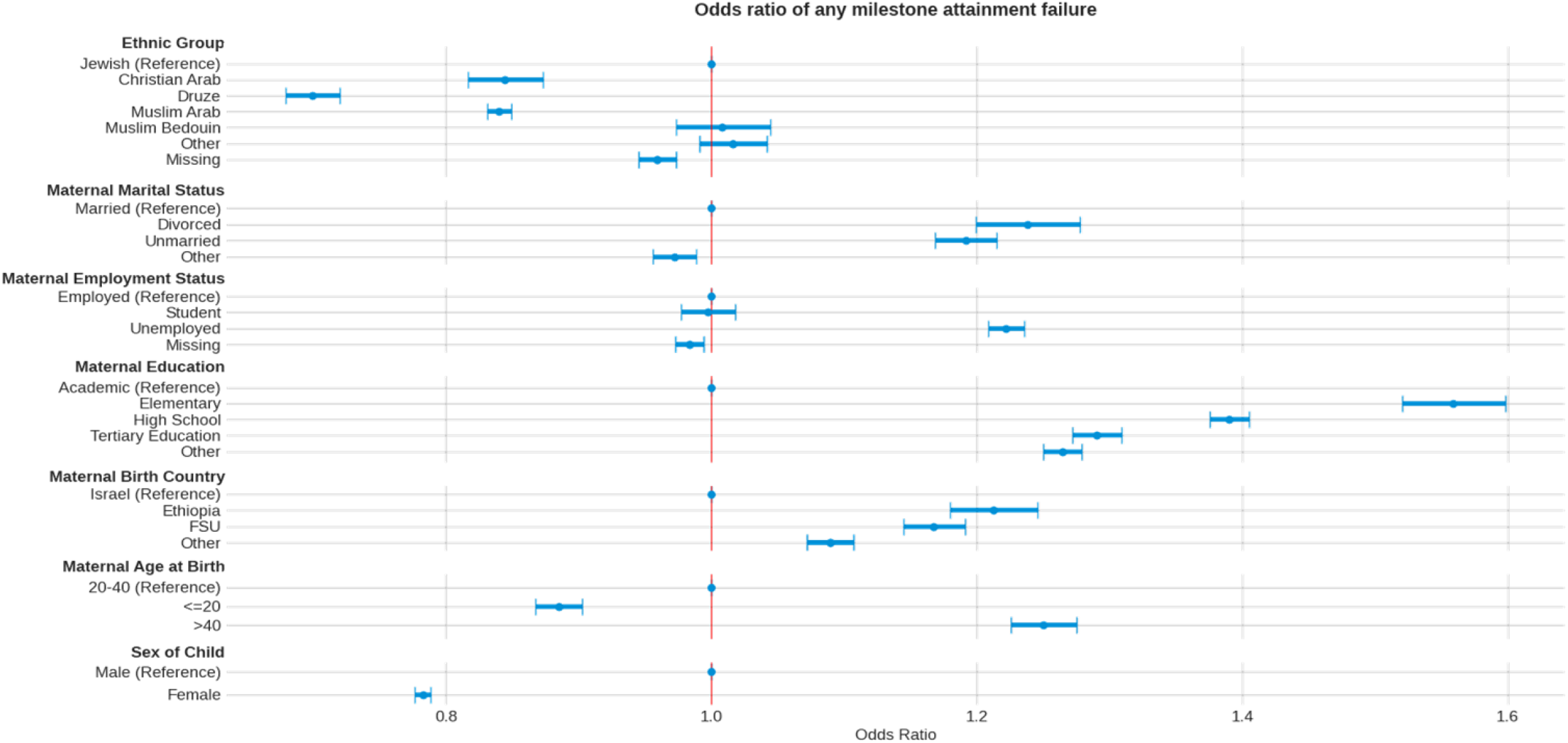
Mutually adjusted odds ratios of any milestone attainment failure, by demographic factors

Higher risk was observed in children to mothers who are divorced (OR=1.24 [1.2, 1.28], P<0.001), or unmarried (OR=1.19 [1.17, 1.21], P<0.001), unemployed (OR=1.22 [1.21, 1.24], P<0.001), immigrant from the FSU (OR=1.17 [1.14, 1.19], P<0.001) or Ethiopia (OR=1.21 [1.18,1.25], P<0.001), have elementary (OR=56 [1.52, 1.6], P<0.001) or high school (OR=1.39 [1.38, 1.41], P<0.001) education or are over 40 years old at time of child’s birth (OR=1.25 [1.23, 1.28], P<0.001). The increased risk was mostly evident in the language, social and fine motor domains.

## Discussion

Attainment of age-dependent milestones is the cornerstone of developmental surveillance tools used for monitoring children worldwide. Here we use a national dataset to describe trends in developmental milestone attainment rates between the years 2016 and 2020, and to evaluate the associations between various sociodemographic factors and these trends.

When evaluating overall rate of failure in attaining any of the milestones, we observed a gradual elevation over the examined years in all developmental domains. This elevation was most prominent in the language domain. These results are consistent with the reported elevation in developmental disorders prevalence over the last decade [2,22].

Parental concern may be a good predictor of developmental delay [23,24]. However, in this study we observed a trend of decline in parental concern for developmental delay, while the rates of failures in milestone attainment increased. These observations should be noted since parental awareness to their child’s difficulties, may encourage suitable interventions. In addition, there is a need to accompany developmental surveillance tools with appropriate parental education regarding the expected attainment of age-dependent milestones by their child.

Multivariable regression analysis enabled to quantify the contribution of the different sociodemographic variables to the risk of failure in attaining milestones. Higher risk was observed in children to mothers who are divorced, unemployed, immigrant, have lower education, are of Bedouin origin or are over 40 years old at time of child’s birth after adjustment for other sociodemographic factors. The increased risk is mostly evident in the language, social and fine motor domains.

Associations between sociodemographic variables and suspected developmental delay were previously reported. These include lower level of maternal education [15,16,19], maternal employment status [17], maternal marital status [16,18] and maternal age [15,16]. Our results are consistent with these reports.

This study has few notable limitations. First, MCHCs that belong to other health maintenance organizations (∼30%) were not included in this study. However, this subset is nationally representative of child’s sex and maternal age, with a minor overrepresentation of the Arab children population [20]. Second, although the MCHCs used a uniform surveillance protocol, its practical implementation may have varied across geographical locations and cultural subgroups. Third, the surveillance visits to the MCHCs in 2020 were irregular due to COVID-19 pandemic, which may have caused specific changes during this period. Another limitation is that while 95% of the children routinely visit the MCHCs until the age of 24 months, these numbers drop significantly at older ages. However, when comparing characteristics of the children who continued visiting MCHCs at older ages to those who visited only at younger children, no significant differences were found [20]. Finally, in this work we used existing demographic factors collected by MCHCs during routine visits. Hopefully in future reports we will be able to include additional variables, such as socioeconomic status, which might help us better describe child development trends.

We hope that this report may support decision makers in allocating resources and choosing appropriate interventions in subpopulations at risk of developmental delays. An annual update of the measured variables will enable objective evaluation of such interventions, in order to maximize their effectiveness.

## Conclusions

This report describes national trends of child development in the gross motor, fine motor, language, and social domains. These trends are important for stakeholders and policy makers, allowing data driven decision making and resource allocation.

We assessed the socioeconomic variables contributing to the risk of failing to attain developmental milestones and identified subpopulations with increased risk.

An annual report, such as the one presented in this work, will allow an objective evaluation of the effectiveness of intervention programs and a continuous calibration of these programs in attempt to maximize the developmental potential of children in Israel and worldwide.

## Supporting information

STROBE cross-sectional study checklist

## Data Availability

All data produced in the present study are available upon reasonable request to the authors

https://kinstitute.org.il/publication_files/israeli-child-development-report/

## List of abbreviations

AAP: American Academy of Pediatrics
CDC: Centers for Disease Control and prevention
EMR: Electronic Medical Record
FSU: Former Soviet Union
MCHC: Maternal child health clinic
MOH: Ministry of Health
OR: Odds Ratio
THIS: Tipat Halav Israel Screening
VIF: Variance Inflation Factor

## Declarations

### Ethics approval and consent to participate

All methods were performed in accordance with the ethical standards as laid down in the Declaration of Helsinki and its later amendments or comparable ethical standards. The study protocol was approved by the Soroka Medical Center institutional review board, approval number MHC-0006-18. An exemption of informed consent was granted by this ethics committee because the data were anonymous.

### Consent for publication

Not applicable.

### Availability of data and materials

The de-identified patient-level data used for this study contains sensitive information and therefore is not available outside the secured research environment of the Israel Ministry of Health. Summary aggregate level data and analysis code for this study can be made available upon reasonable request to the corresponding author.

## Authors’ contributions

IGi, GA, DRZ, RB, PA, MAT and YS conceptualized and designed the study. IGi, GA, IGo, DRZ, PA, MAT and YS contributed to data acquisition, analysis, or interpretation. IGi, GA and YS drafted the manuscript. IGi, GA, IGo, DRZ, RB, PA, MAT and YS revised the manuscript. IGi and IGo performed statistical analysis. DRZ, RB and MAT supplied technical or administrative support for the study. All authors read and approved the final manuscript.

## Acknowledgements

This study was conducted with the help of TIMNA – a national research platform established by the Israel government to enable big-data studies combining de-identified health data from multiple organizations.

## Supplementary figures

**Supplementary Figure 1.**
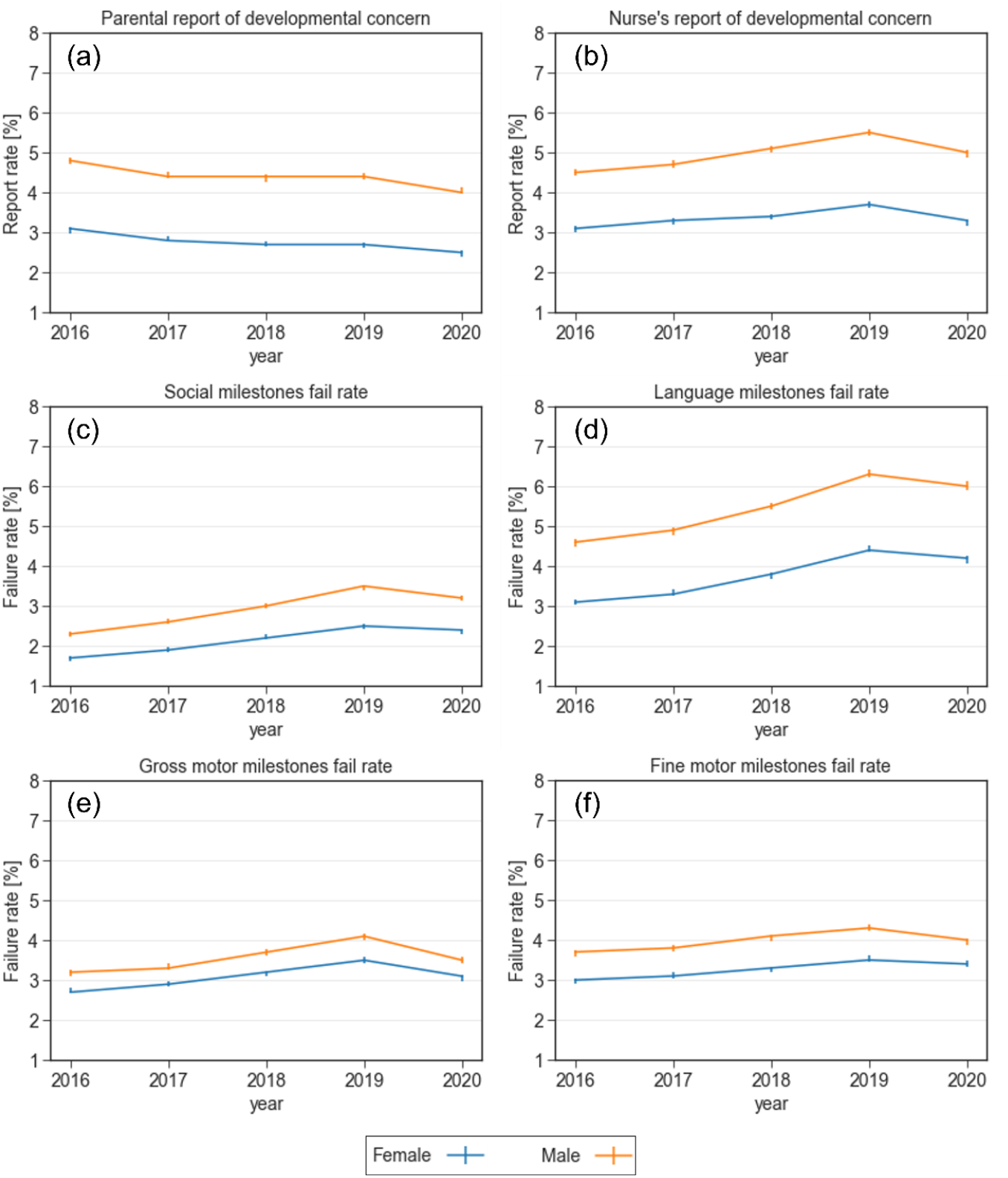
Reports of concern and failure rates in various developmental domains stratified by child’s sex between 2016-2020. (a) Parental report of developmental concern (b) Nurse’s report of developmental concern (c) Failure rate in the social domain (d) Failure rate in the language domain (e) Failure rate in the gross motor domain (f) Failure rate in the fine motor domain

**Supplementary Figure 2.**
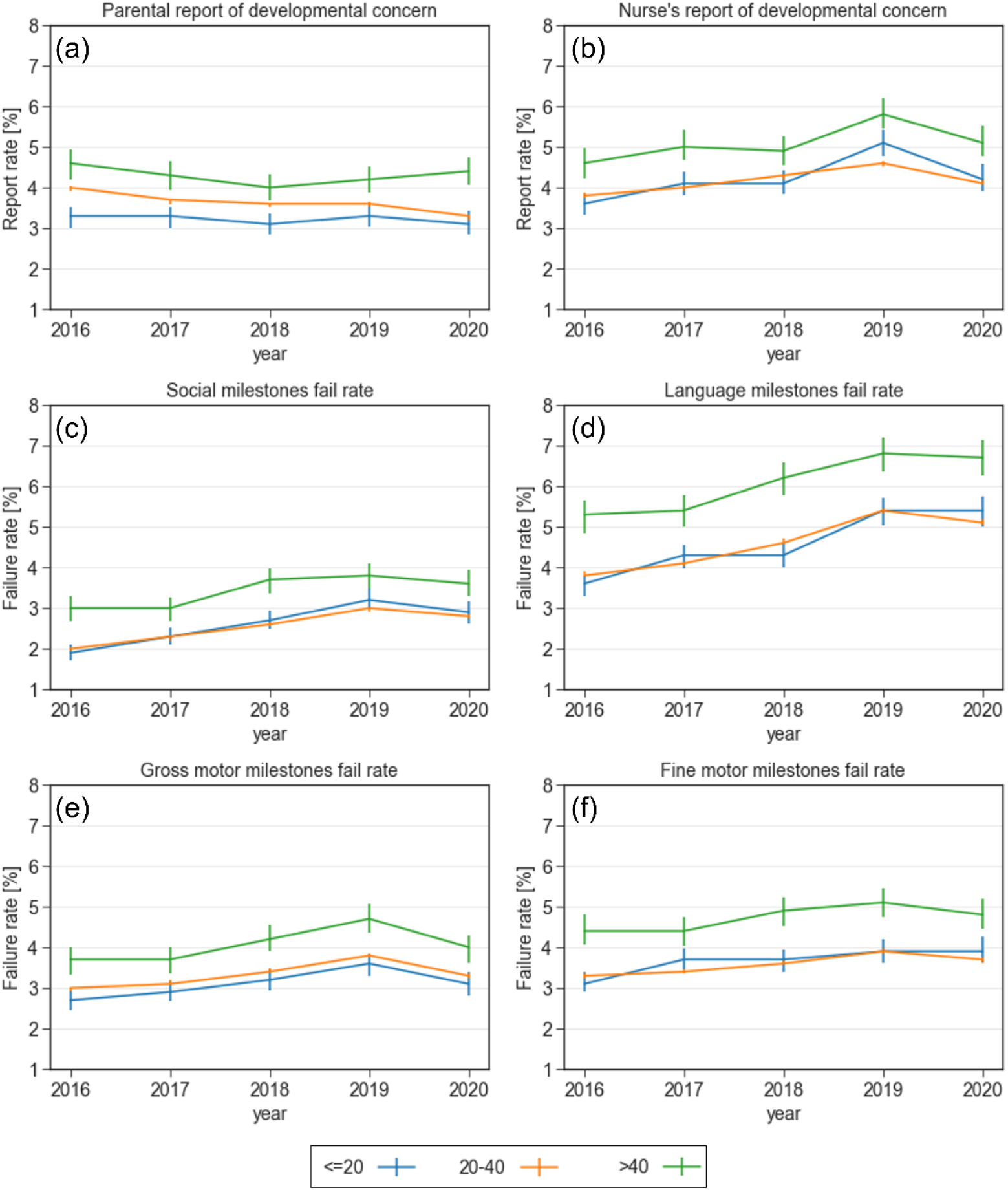
Reports of concern and failure rates in various developmental domains stratified by maternal age between 2016-2020. (a) Parental report of developmental concern (b) Nurse’s report of developmental concern (c) Failure rate in the social domain (d) Failure rate in the language domain (e) Failure rate in the gross motor domain (f) Failure rate in the fine motor domain

**Supplementary Figure 3.**
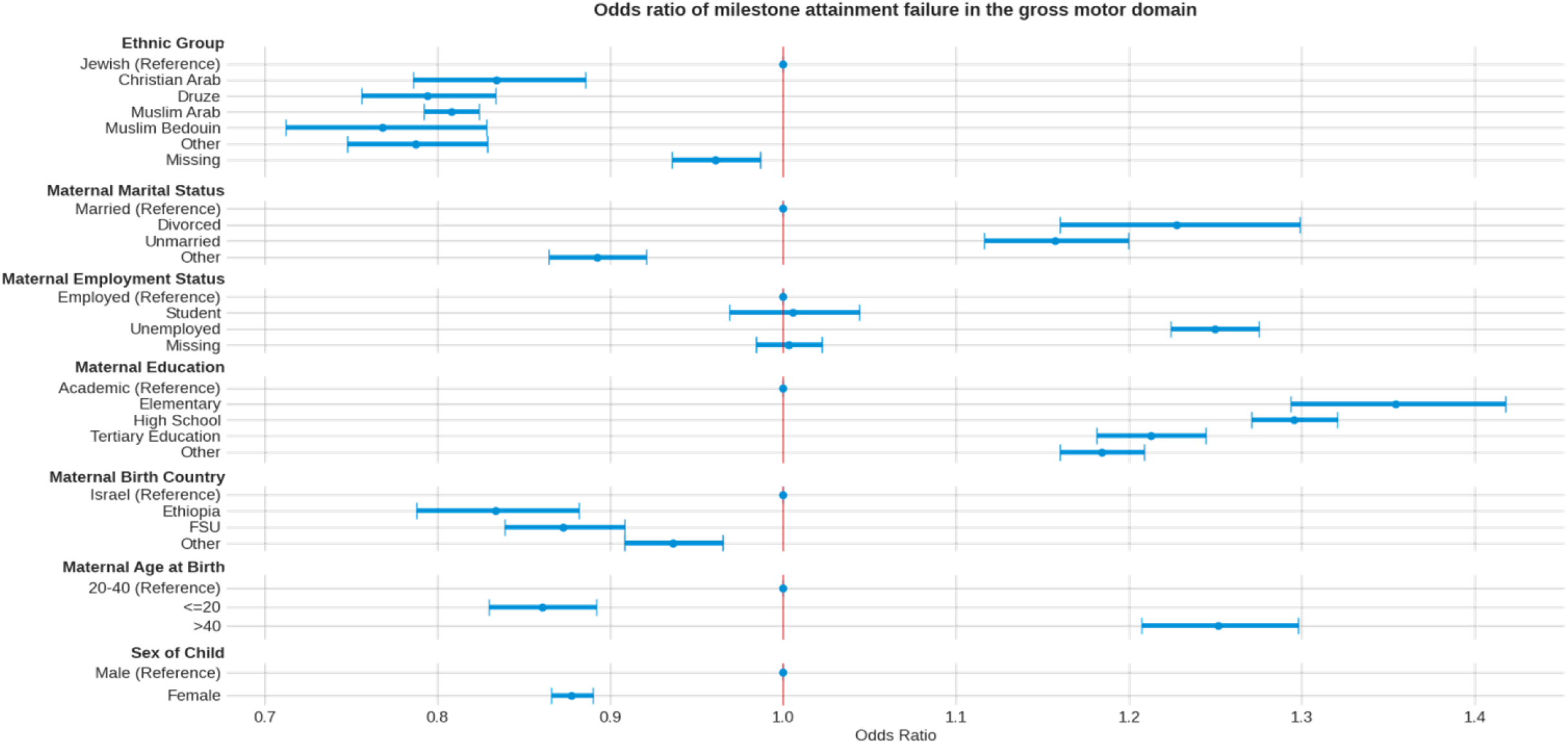
Mutually adjusted odds ratios of milestone attainment failure in the gross motor domain, by demographic factors

**Supplementary Figure 4.**
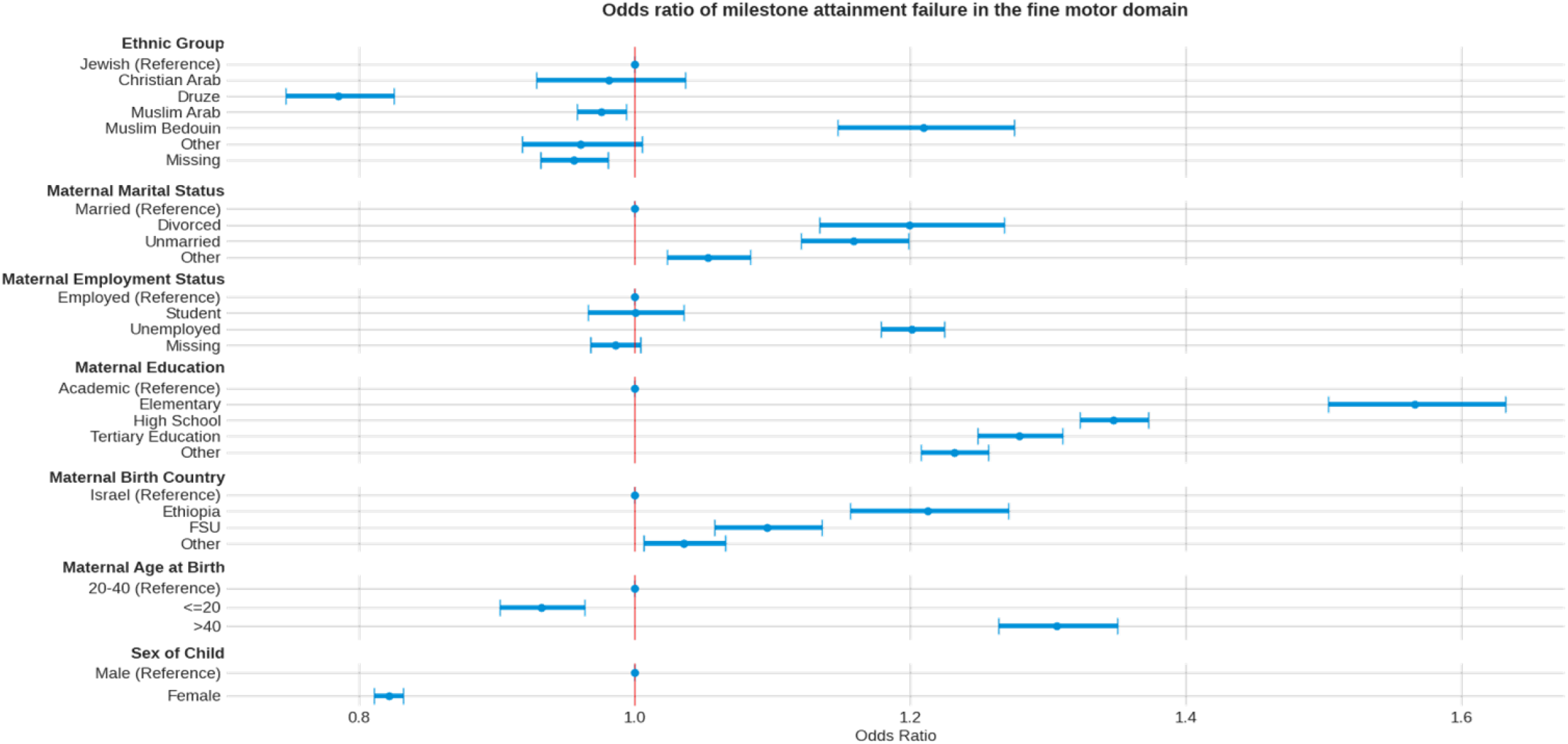
Mutually adjusted odds ratios of milestone attainment failure in the fine motor domain, by demographic factors

**Supplementary Figure 5.**
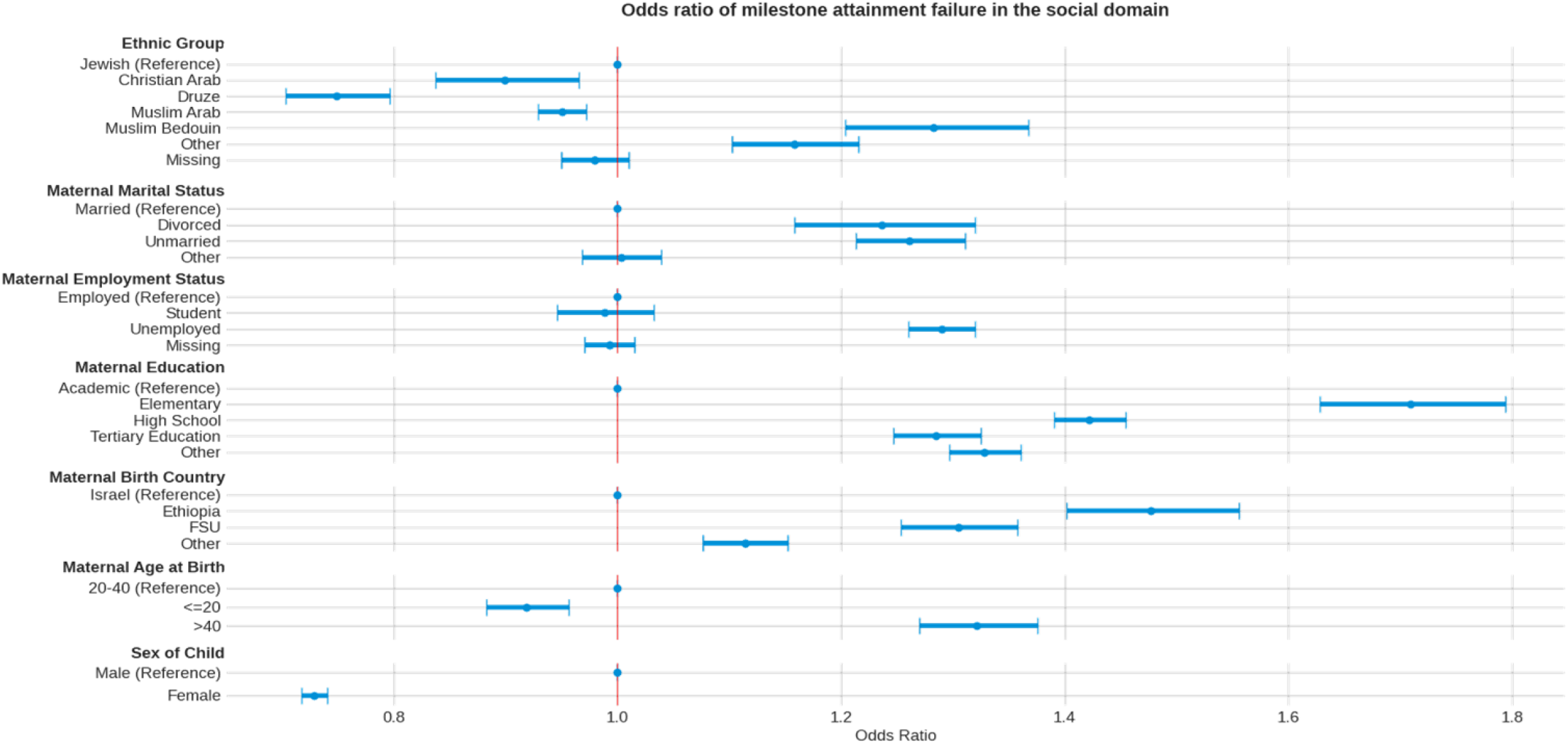
Mutually adjusted odds ratios of milestone attainment failure in the social domain, by demographic factors

**Supplementary Figure 6.**
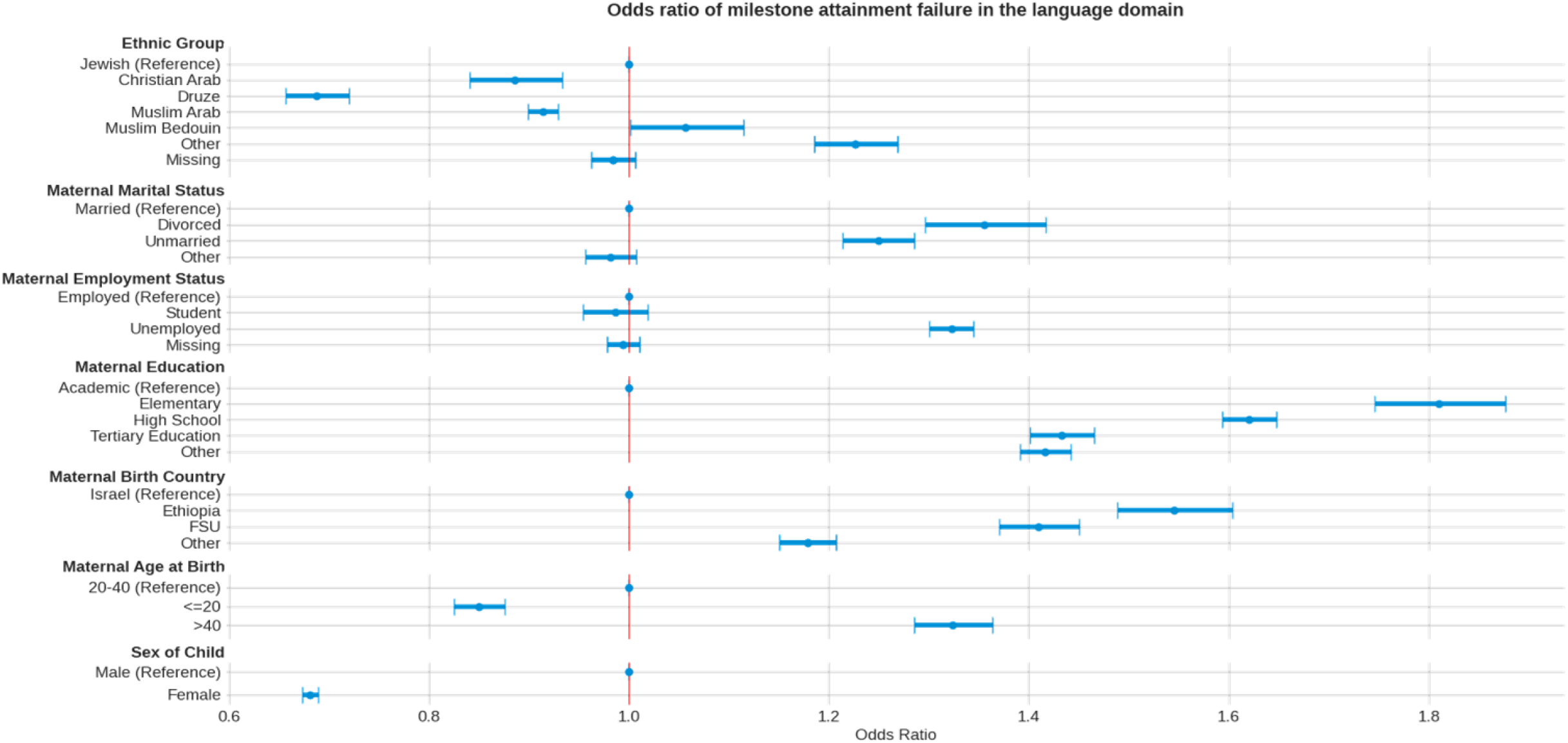
Mutually adjusted odds ratios of milestone attainment failure in the language domain, by demographic factors

